# Genome-wide meta-analysis identifies novel maternal risk variants and enables polygenic prediction of preeclampsia and gestational hypertension

**DOI:** 10.1101/2022.11.30.22282929

**Authors:** Michael C. Honigberg, Buu Truong, Raiyan R. Khan, Brenda Xiao, Laxmi Bhatta, Thi Ha Vy, Rafael F. Guerrero, Art Schuermans, Margaret Sunitha Selvaraj, Aniruddh P. Patel, Satoshi Koyama, So Mi Jemma Cho, Shamsudheen Karuthedath Vellarikkal, Mark Trinder, Sarah M. Urbut, Kathryn J. Gray, Ben M. Brumpton, Snehal Patil, Sebastian Zöllner, Mariah C. Antopia, Genes & Health Research Team, Estonian Biobank Research Team, nuMoM2b Research Team, Richa Saxena, Girish N. Nadkarni, Ron Do, Qi Yan, Itsik Pe’er, Shefali Setia Verma, Rajat M. Gupta, David M. Haas, Hilary C. Martin, David A. van Heel, Triin Laisk, Pradeep Natarajan

**Affiliations:** Cardiology Division, Department of Medicine, Massachusetts General Hospital, Boston, MA, USA; Cardiovascular Research Center, Massachusetts General Hospital, Harvard Medical School, Boston, MA, USA; Program in Medical and Population Genetics, Broad Institute of Harvard and MIT, Cambridge, MA, USA; Department of Computer Science, Columbia University, New York, NY, USA; University of Pennsylvania, Philadelphia, PA, USA; K.G. Jebsen Center for Genetic Epidemiology, Department of Public Health and Nursing, NTNU, Norwegian University of Science and Technology, Trondheim, Norway; HUNT Research Centre, Department of Public Health and Nursing, NTNU, Norwegian University of Science and Technology, Levanger, Norway; The Charles Bronfman Institute for Personalized Medicine, Icahn School of Medicine at Mount Sinai, New York, NY, USA; Department of Biological Sciences, North Carolina State University, Raleigh, NC, USA; Faculty of Medicine, KU Leuven, Leuven, Belgium; Integrative Research Center for Cerebrovascular and Cardiovascular Diseases, Seoul, Republic of Korea; Cardiovascular Division, Department of Medicine, Brigham and Women’s Hospital, Boston, MA, USA; Centre for Heart Lung Innovation, University of British Columbia, Vancouver, BC, Canada; Department of Obstetrics and Gynecology, Brigham and Women’s Hospital, Boston, MA, USA; Center for Genomic Medicine, Massachusetts General Hospital, Boston, MA, USA; Department of Biostatistics and Center for Statistical Genetics, University of Michigan School of Public Health, Ann Arbor, MI, USA; Department of Integrative Biology, University of Texas at San Antonio, San Antonio, TX, USA; Department of Obstetrics and Gynecology, Columbia University, New York, NY, USA; Indiana University School of Medicine, Indianapolis, IN, USA; Department of Human Genetics, Wellcome Sanger Institute, Cambridge, UK; Blizard Institute, Barts and The London School of Medicine and Dentistry, Queen Mary University of London, London, UK; Estonian Genome Centre, Institute of Genomics, University of Tartu, Tartu, Estonia

## Abstract

Preeclampsia and gestational hypertension are common pregnancy complications associated with adverse maternal and offspring outcomes. Current tools for prediction, prevention, and treatment are limited. We tested the association of maternal DNA sequence variants with preeclampsia in 20,064 cases and 703,117 controls and with gestational hypertension in 11,027 cases and 412,788 controls across discovery and follow-up cohorts using multi-ancestry meta-analysis. Altogether, we identified 18 independent loci associated with preeclampsia/eclampsia and/or gestational hypertension, 12 of which are novel (e.g., *MTHFR-CLCN6*, *WNT3A*, *NPR3*, *PGR*, and *RGL3*), including two loci (*PLCE1*, *FURIN*) identified in multi-trait analysis. Identified loci highlight the role of natriuretic peptide signaling, angiogenesis, renal glomerular function, trophoblast development, and immune dysregulation. We derived genome-wide polygenic risk scores that predicted preeclampsia/eclampsia and gestational hypertension in external datasets, independent of first trimester risk markers. Collectively, these findings provide mechanistic insights into the hypertensive disorders of pregnancy and advance pregnancy risk stratification.

## Introduction

The hypertensive disorders of pregnancy (HDPs) represent a leading global cause of maternal and neonatal morbidity and mortality and account for approximately 14% of maternal deaths worldwide^1,2^. Up to 15% of child-bearing women experience an HDP in at least one pregnancy^3,4^. The HDPs include preeclampsia, defined as new-onset hypertension or worsening hypertension after 20 weeks’ gestation plus proteinuria or other evidence of end-organ dysfunction; gestational hypertension, defined as new-onset hypertension without accompanying features of preeclampsia; and eclampsia, defined as progression of preeclampsia to maternal seizures^5,6^. In addition to short-term risks of end-organ failure and death in the absence of prompt recognition and treatment, those who develop HDPs have roughly two-fold long-term risk of cardiovascular disease events and premature cardiovascular mortality compared with those who experience only normotensive pregnancies for reasons that remain incompletely understood^7,8^.

The pathophysiology of the HDPs is increasingly recognized to be heterogeneous with both maternal and fetal contributions. In the contemporary model of preeclampsia pathophysiology, defective trophoblast invasion in early placental development and incomplete remodeling of the maternal spiral arteries lead to placental ischemia later in gestation^1,9,10^. The distressed placenta secretes an excess of circulating anti-angiogenic proteins (e.g., soluble fms-like tyrosine kinase receptor 1 [sFlt-1] and soluble endoglin) which induce the systemic maternal endothelial dysfunction and vasoconstriction that drive the clinical manifestations of preeclampsia (hypertension and proteinuria)^11^. In addition, maternal cardiometabolic risk factors (e.g., pre-pregnancy chronic hypertension, diabetes, obesity) and pre-pregnancy kidney and autoimmune disease strongly predict preeclampsia^1,12^ and influence early placentation as well as maternal vascular adaptation to pregnancy^10,13^.

Genetic analysis may yield novel mechanistic insights into the pathophysiology of the HDPs and may also help improve pregnancy risk prediction. An estimated 31-35% of preeclampsia predisposition has been attributed to maternal genetics using familial aggregation-based approaches^14,15^. However, few genetic loci linked to preeclampsia have been identified and robustly validated to date. Several fetal variants near the *FLT1* gene, which encodes placenta-derived sFlt-1, have been reported to associate with preeclampsia^16,17^. Steinthorsdottir et al. recently performed the largest maternal GWAS of preeclampsia to date and identified associations near *FTO* (the first reported obesity-associated locus)^18^, *ZNF831*, and several other blood pressure-associated genes (*MECOM*, *FGF5*, and *SH2B3*) in combined meta-analysis of 12,150 cases and 164,098 controls^17^. In addition, several studies have now reported that increased maternal polygenic risk of hypertension associates with risk of HDPs^17,19–22^.

In this work, we performed expanded multi-ancestry GWAS meta-analysis for preeclampsia/eclampsia and separately performed GWAS for gestational hypertension. We then used these results to train and test polygenic risk scores (PRS) for each outcome in independent datasets (**Extended Data Fig. 1**).

## Results

### Association of common genetic variants with preeclampsia/eclampsia and gestational hypertension

We tested the association of common variants (minor allele frequency [MAF] ≥1%) with preeclampsia/eclampsia among 17,150 cases and 451,241 controls in discovery analysis (78.0% European, 21.2% Asian, 0.5% admixed American, and 0.3% African ancestry; **Supplementary Table 1**) using multi-ancestry fixed-effects meta-analysis in METAL^23^. Cases were identified principally using *International Classification of Diseases* (ICD) codes and phecodes corresponding to preeclampsia and, where available, eclampsia (**Supplementary Tables 1-3**); controls were generally either women with exclusively normotensive pregnancies or all women without codes corresponding to hypertension in pregnancy^17^. In discovery analysis, we identified 12 independent loci at the commonly used statistical significance threshold of *P*<5×10^−8^, including 6 previously nominated by Steinthorsdottir et al. in maternal or fetal GWAS (*MECOM* [3q26], *FGF5* [4q21], *SH2B3* [12q22], *FLT1* [13q12], *FTO* [16q12], and *ZNF831* [20q13])^17^ and 6 additional loci (*MTHFR-CLCN6* [1p36], *WNT3A* [1q42], *MICA* [6p21], *LINC00484* [9q22], *PGR* [11q22], and *RGL3* [19p13]; **Table 1, Supplementary Fig. 2a, Supplementary Table 4**).

**Table 1.** Maternal sequence variants associated with preeclampsia/eclampsia.

|  |  |  |  |  | Discovery analysis<br>(17,150 cases / 451,241 controls) |  |  |  | Follow-up analysis<br>(2,914 cases / 251,876 controls) |  | Combined discovery and follow-up meta-analysis<br>(20,064 cases / 703,117 controls) |  |
| --- | --- | --- | --- | --- | --- | --- | --- | --- | --- | --- | --- | --- |
| Nearest gene | Lead variant | CHR | POS | Effect allele / other allele | Weighted average EAF | OR | P-value | P(het)* | OR | P-value | OR | P-value |
| *Loci with genome-wide significance in discovery analysis* |  |  |  |  |  |  |  |  |  |  |  |  |
| *MTHFR / CLCN6* | rs149764880 | 1 | 11820674 | G / T | 0.85 | 1.11 | 1.8e-8 | 0.30 | 1.17 | 6.8e-4 | 1.12 | **8.8e-11** |
| *WNT3A* | rs708119 | 1 | 228015567 | C / G | 0.67 | 1.08 | 2.3e-8 | 0.92 | 1.05 | 0.10 | 1.08 | **7.8e-9** |
| *MECOM* | rs9855086 | 3 | 169413542 | A / T | 0.53 | 1.08 | 5.7e-10 | 0.21 | 1.08 | 8.3e-3 | 1.08 | **1.6e-11** |
| *FGF5* | rs16998073 | 4 | 80263187 | T / A | 0.32 | 1.11 | 3.1e-14 | 0.15 | 1.08 | 1.6e-2 | 1.11 | **2.3e-15** |
| *MICA* | rs2442752 | 6 | 31383987 | C / T | 0.36 | 1.08 | 2.3e-9 | 0.57 | 0.99 | 0.84 | 1.07 | **3.8e-8** |
| *LINC00484* | rs5899121 | 9 | 91145498 | TG / T | 0.85 | 1.12 | 4.6e-8 | 0.31 | 0.86 | 0.31 | 1.12 | 1.4e-7 |
| *PGR* | rs2508372 | 11 | 101397592 | A / G | 0.79 | 1.10 | 1.3e-9 | 0.14 | 1.11 | 7.5e-3 | 1.11 | **3.4e-11** |
| *SH2B3* | rs10774624 | 12 | 111395984 | G / A | 0.47 | 1.10 | 2.9e-12 | 0.40 | 1.03 | 0.29 | 1.09 | **1.0e-11** |
| *FLT1* | rs7318880 | 13 | 28564148 | T / C | 0.48 | 1.09 | 6.7e-12 | 0.79 | 1.06 | 5.8e-2 | 1.09 | **1.6e-12** |
| *FTO* | rs1421085 | 16 | 53767042 | C / T | 0.40 | 1.10 | 4.7e-12 | 0.61 | 1.07 | 1.8e-2 | 1.09 | **3.3e-13** |
| *RGL3* | rs167479 | 19 | 11416089 | G / T | 0.55 | 1.10 | 7.6e-10 | 0.20 | 1.09 | 6.8e-3 | 1.10 | **1.8e-11** |
| *ZNF831* | rs259983 | 20 | 59160402 | C / A | 0.16 | 1.13 | 1.0e-12 | 0.40 | 1.15 | 2.8e-4 | 1.14 | **1.3e-15** |
| *Additional loci identified in combined discovery and follow-up meta-analysis* |  |  |  |  |  |  |  |  |  |  |  |  |
| *FGL1* | rs2653414 | 8 | 17868560 | A / C | 0.02 | 1.27 | 1.0e-6 | 0.44 | 2.18 | 4.4e-9 | 1.36 | **3.4e-11** |
| *UPB1* | rs17572606 | 22 | 24472204 | T / C | 0.02 | 1.45 | 1.9e-7 | 0.33 | 1.28 | 6.0e-2 | 1.41 | **4.6e-8** |

\*P(het) indicates P-value for heterogeneity across discovery cohorts.
CHR indicates chromosome. POS indicates position (genome build 38). EAF indicates effect allele frequency. OR indicates odds ratio.

We pursued replication of these GWAS results in 4 follow-up cohorts that collectively included 2,914 preeclampsia/eclampsia cases and 251,876 controls (96.7% European, 3.1% African, and 0.3% admixed American ancestry). We replicated 7 of 12 associations from discovery analysis with *P*<0.05 and consistent direction of effect, including the novel associations at *MTHFR-CLCN6*, *PGR*, and *RGL3* (**Table 1**). Ten associations had consistent direction of effect in follow-up cohorts, and 11 of 12 associated loci retained genome-wide significance in combined meta-analysis of preeclampsia/eclampsia discovery and follow-up cohorts. In meta-analysis of discovery and follow-up cohorts, 2 additional loci attained genome-wide significance (*FGL1* [8p22] and *UPB1* [22q11]), yielding a total of 13 loci associated with preeclampsia/eclampsia with genome-wide significance in combined meta-analysis (**Fig. 1a**). We did not observe inflation in test statistics (lambda genomic inflation factor: 1.038, **Supplementary Fig. 1a**). There was no discernible heterogeneity of these associations across ancestries (**Supplementary Table 5**). Conditional analysis in GCTA-COJO^24^ identified a second independent association on chromosome 20 near *ZBTB46* (lead variant rs4809370, OR 1.08, *P*=1.4×10^−8^).

**Figure 1.**
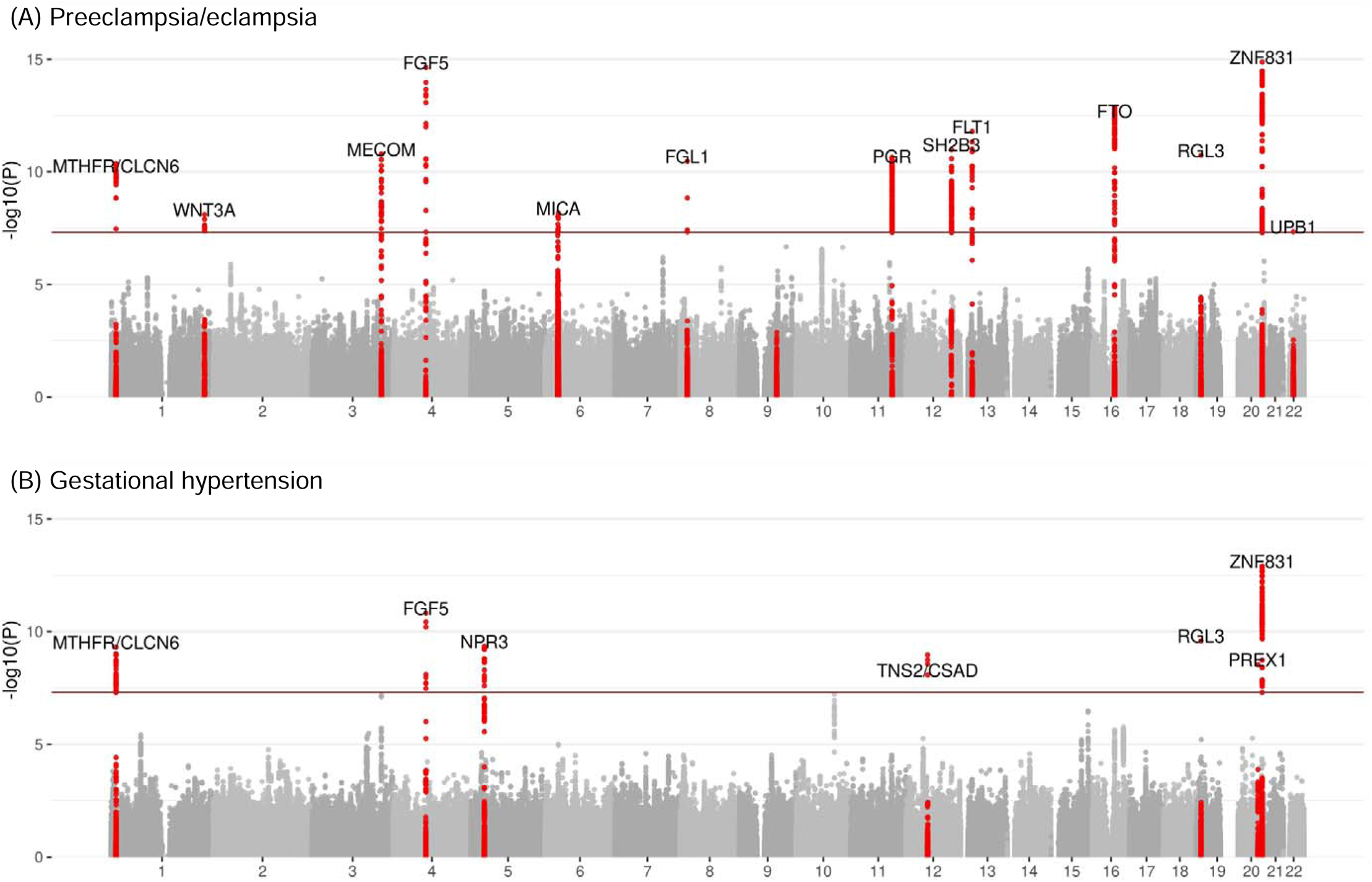
Manhattan plots of preeclampsia/eclampsia and gestational hypertension in combined discovery and follow-up meta-analysis. Manhattan plots (chromosomal position on the X-axis and −log(10) of the P-value on the Y-axis) are displayed for (A) preeclampsia/eclampsia in 20,064 cases and 703,117 controls and (B) gestational hypertension in 11,027 cases and 412,788 controls. Analyses included multi-ancestry meta-analysis of common variants (minor allele frequency ≥1%). Loci are labeled by the gene nearest to the lead variant.

We next tested the association of common variants with gestational hypertension among 8,961 cases and 184,925 controls in discovery analysis (91.3% European, 6.7% Asian, 0.7% African, 1.3% admixed American) and among 2,066 cases and 227,863 controls in follow-up cohorts (96.3% European, 3.4% African, 0.3% admixed American, **Supplementary Tables 1-2**). Gestational hypertension cases were identified primarily based on qualifying ICD codes for gestational hypertension and an absence of qualifying codes for preeclampsia/eclampsia. In discovery analysis, we identified 7 independent genome-wide significant loci associated with gestational hypertension, including 4 also associated with preeclampsia/eclampsia (*MECOM*, *FGF5*, *RGL3*, and *ZNF831*) and 3 additional associations (*NPR3* [5p13], *TNS2-CSAD* [12q13], and *PREX1* [20q13]; **Table 2**). Four of 7 significant associations replicated with *P*<0.05 in follow-up cohorts (*FGF5*, *RGL3*, *PREX1*, and *ZNF831*), and all 7 loci had consistent direction of effect in follow-up cohorts. In combined meta-analysis of discovery and follow-up cohorts, 6 of 7 loci retained genome-wide significance, and the *MTHFR-CLCN6* locus (1p36) additionally reached genome-wide significance, yielding a total of 7 loci associated with gestational hypertension in combined meta-analysis (**Fig. 1b**). As with preeclampsia/eclampsia, we did not observe inflation in test statistics (lambda genomic inflation factor: 0.976, **Supplementary Fig. 1b**). Stratified analyses suggested potential heterogeneity of association by ancestry (*P*_heterogeneity_=0.001) at the *MECOM* locus, with an inverse association with the lead risk variant observed in those with admixed American ancestry (**Supplementary Table 5**).

**Table 2.**
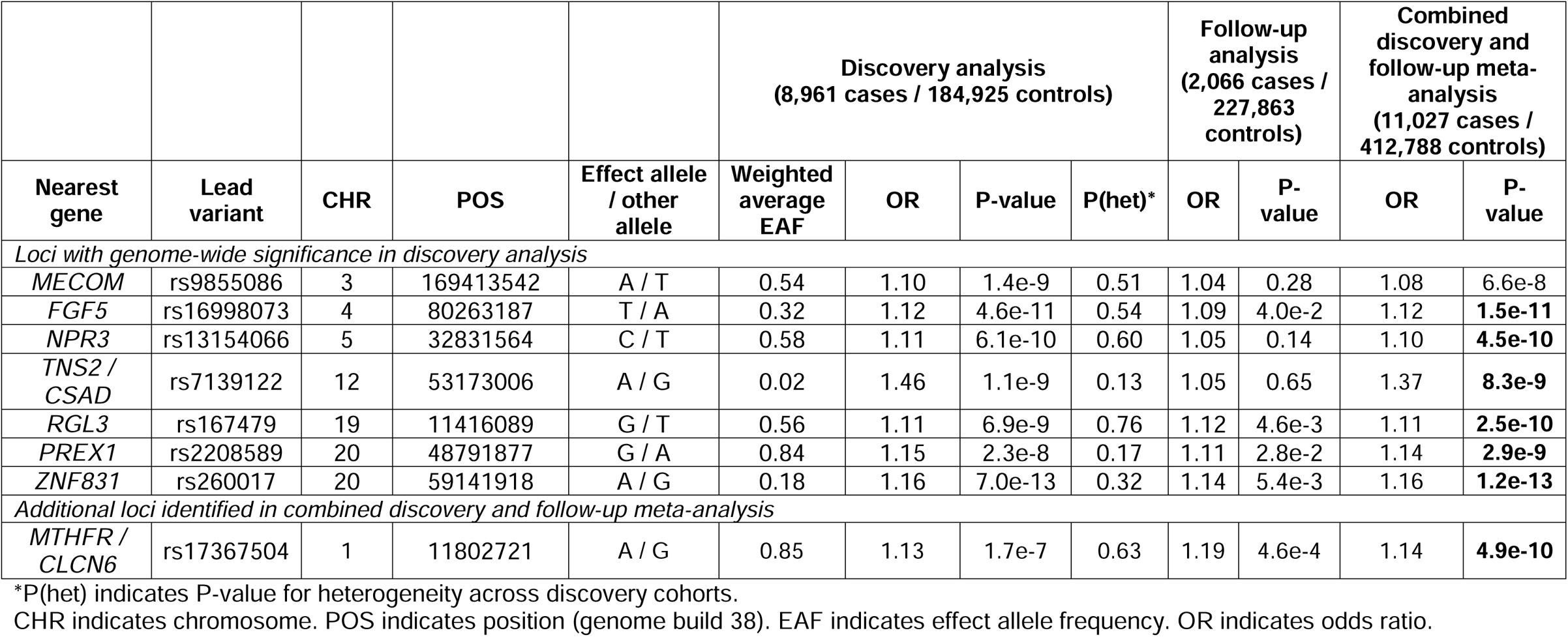
Maternal sequence variants associated with gestational hypertension.

### Genetic correlation across hypertension-related phenotypes

We used cross-trait LD score regression^25^ to assess genetic correlations among preeclampsia/eclampsia, gestational hypertension, systolic blood pressure (SBP), and diastolic blood pressure (DBP)^26^. Preeclampsia/eclampsia and gestational hypertension were strongly genetically correlated (*r*_g_ = 0.71, SE = 0.08). SBP demonstrated stronger genetic correlation with gestational hypertension (*r*_g_ = 0.73, SE = 0.06) vs. preeclampsia/eclampsia (*r*_g_ = 0.52, SE = 0.05). Of note, correlation of SBP with gestational hypertension (*r*_g_ = 0.73) was larger than that of SBP with DBP (*r*_g_ = 0.62, SE = 0.03, in the Million Veteran Program^26^, with similar genetic correlation between SBP and DBP observed previously in the UK Biobank^27^). Genetic correlations with the HDPs were stronger for SBP vs. DBP (**Supplementary Table 6**).

### Multi-trait analysis of GWAS

Given the high degree of genetic correlation observed between preeclampsia/eclampsia and gestational hypertension noted earlier, we used multi-trait analysis of genome-wide association summary statistics (MTAG)^28^ to boost power to identify additional associated variants. Consistent with this high degree of correlation, MTAG yielded very similar results for each trait (displayed for preeclampsia/eclampsia in **Extended Data Fig. 2**). MTAG identified 2 additional loci with genome-wide significance (**Supplementary Table 7**): *PLCE1* (10q23), a blood pressure-associated gene involved in glomerular podocyte development^29^, which also narrowly missed statistical significance in combined meta-analysis of gestational hypertension discovery and follow-up cohorts (*P*=6.0×10^−8^); and *FURIN* (15q26), whose product is a protein convertase involved in processing pro-natriuretic peptides^30^ and whose expression is decreased in preeclamptic placentas^31^.

### Gene prioritization at preeclampsia/eclampsia and gestational hypertension risk loci

To prioritize causal genes, we first performed colocalization analysis with expression quantitative trait loci (eQTLs) within ±500 kilobases of lead variants across 52 tissues in the Genotype-Tissue Expression project (GTEx) (**Supplementary Tables 8-9**)^32^. Colocalization analyses implicated *FGF5* and *NPR3* as causal genes at their respective loci. The lead variant at the *MTHFR-CLCN6* locus colocalized with *CLCN6* eQTLs as well as expression of *NPPA* (which encodes the precursor to atrial natriuretic peptide [ANP]). The lead preeclampsia/eclampsia variant at the *ZNF831* locus colocalized with multiple genes but most strongly with *ZBTB46* expression, including in arterial tissue. We also observed multiple colocalizations with *WNT3A* (*WNT3A*, *GJC2,* and mitochondrial proteins *IBA57* and *MPRL55*), *MICA* (*CLIC1* and psoriasis-associated genes *TCF19*, *CCHCR1*, and *PSORS1C1*), and *RGL3* (*ZNF627* and *EPOR*). We observed no strong colocalizations with lead variants at *LINC00484*, *PGR*, *SH2B3*, *FLT1*, *FTO*, or *TNS2-CSAD*.

Next, we queried variant-to-gene evidence in Open Targets Genetics v7 (**Supplementary Tables 10-11**)^33^ and generated polygenic priority scores (PoPS, **Supplementary Table 12**)^34^ for lead variants. Both approaches nominated *MECOM*, *FGF5*, *SH2B3*, *FTO*, and *NPR3* as the most likely causal gene and their respective loci. PoPS prioritized *NPPA* as the most likely causal gene at the *MTHFR-CLCN6* locus and *TRPC6* as the most likely causal gene at the *PGR* locus.

To further understand how identified genes might influence HDP risk, we queried lead maternal variants in the fetal GWAS for maternal preeclampsia from Steinthorsdottir et al.^17^ (**Supplementary Table 13**). As published previously^16,17^, the lead *FLT1* variant was strongly associated with preeclampsia (*P*=3.9×10^−11^); all other lead variants had *P*>10^−4^ in the fetal GWAS. In addition, we queried nearest genes and those prioritized by colocalization, variant-to-gene scores, and/or PoPS in a publicly available database of the human placental transcriptome including preeclampsia cases and controls (**Supplementary Table 14**)^35^. Consistent with the correlation of increased circulating placental sFlt-1 with preeclampsia incidence^11,36^, *FLT1* gene expression was increased in preeclamptic placentas (log_2_(fold change) 0.39, false discovery rate-adjusted *P*=0.003). Expression of *WNT3A,* which occurs almost exclusively in the placenta^37^, was increased in preeclamptic placentas vs. healthy controls (log_2_(fold change) 0.21, adjusted *P*=0.029), in line with another recent study^38^. *OBSCN,* which was most strongly prioritized by PoPS at the *WNT3A* locus, was also overexpressed in preeclamptic vs. control placentas (log_2_(fold change) 0.18, adjusted *P*=0.037). Furthermore, preeclamptic placentas demonstrated lower expression of *ARHGAP42*—which sits adjacent to *PGR* and encodes Rho GTP-ase activating protein 42, a known regulator of vascular tone and blood pressure expressed selectively in smooth muscle cells^39^—compared with controls (log_2_(fold change) −0.18, adjusted *P*=0.004).

We analyzed expression of prioritized genes in a dataset of single-nuclei RNA sequencing (snRNA-seq) from non-atherosclerotic human aortic tissue. SnRNA-seq identified subpopulations of vascular smooth muscle cells, fibromyocytes, fibroblasts, endothelial cells, macrophages, natural killer T cells, and neuronal cells. The greatest enrichment was seen in the two endothelial cell (EC) populations and in macrophages (**Extended Data Fig. 3**). The EC1 subpopulation is enriched for genes in angiogenesis and lipoprotein assembly and clearance, while the EC2 subpopulation is enriched for genes in extracellular matrix production and integrin expression^40^. Relative expression in ECs vs. other cell types was strongest for *FLT1* (EC1/EC2), *ZBTB46* (EC1), and *MECOM* (EC2).

### Training and testing polygenic risk scores for preeclampsia/eclampsia and gestational hypertension

We used PRS-CS^41^ to construct genome-wide PRS for preeclampsia/eclampsia (PRS_preeclampsia_) and gestational hypertension (PRS_GH_) from our corresponding discovery GWAS summary statistics. In addition, since blood pressure polygenic risk has previously been associated with HDPs^17,19,20^, we used PRS-CS to derive a PRS for SBP (PRS_SBP_) using the SBP GWAS from the Million Veteran Program^26^ to compare prediction across scores and determine whether a linear combination of each HDP PRS and PRS_SBP_ improves performance.

We tuned polygenic scores among 423 women with a history of preeclampsia/eclampsia, 482 women with a history of gestational hypertension only, and 211,129 parous women without a reported history of HDPs in the UK Biobank. The global shrinkage parameter of 1×10^−4^ was chosen for PRS_preeclampsia_ and PRS_SBP_ and of 1×10^−6^ for PRS_GH_ as these values generated the highest *R*^2^ (**Supplementary Table 15**). A linear combination of PRS_preeclampsia_ and PRS_SBP_ (PRS_preeclampsia+SBP_) improved performance vs. each score individually for the outcome of preeclampsia (**Supplementary Table 16**).

The individual and combination PRS tuned in the UK Biobank were carried forward for external validation in two complementary datasets: a Norwegian population-based cohort linked to the Medical Birth Register of Norway (HUNT, preeclampsia/eclampsia only) and a prospective U.S. pregnancy cohort (nuMoM2b). Among 25,582 Norwegian women in HUNT (1,569 [6.1%] with preeclampsia/eclampsia), the prevalence of preeclampsia/eclampsia ranged from ~4% among women in the bottom decile of PRS_preeclampsia+SBP_ to ~10% among the top decile of PRS_preeclampsia+SBP_ (**Fig. 2a**). After adjustment for age at birth, age^2^, and the first 10 principal components (PC) of ancestry, the OR corresponding to the top 10%vs. bottom 90% of PRS_preeclampsia+SBP_ was 1.85 (95% CI 1.61-2.13, *P*=6.3×10^−18^). PRS_preeclampsia+SBP_ increased Nagelkerke’s *R*^2^ by 28.5% compared with the PRS_preeclampsia_ alone and by 79.3% compared with PRS_SBP_ alone (**Supplementary Table 17**).

**Figure 2.**
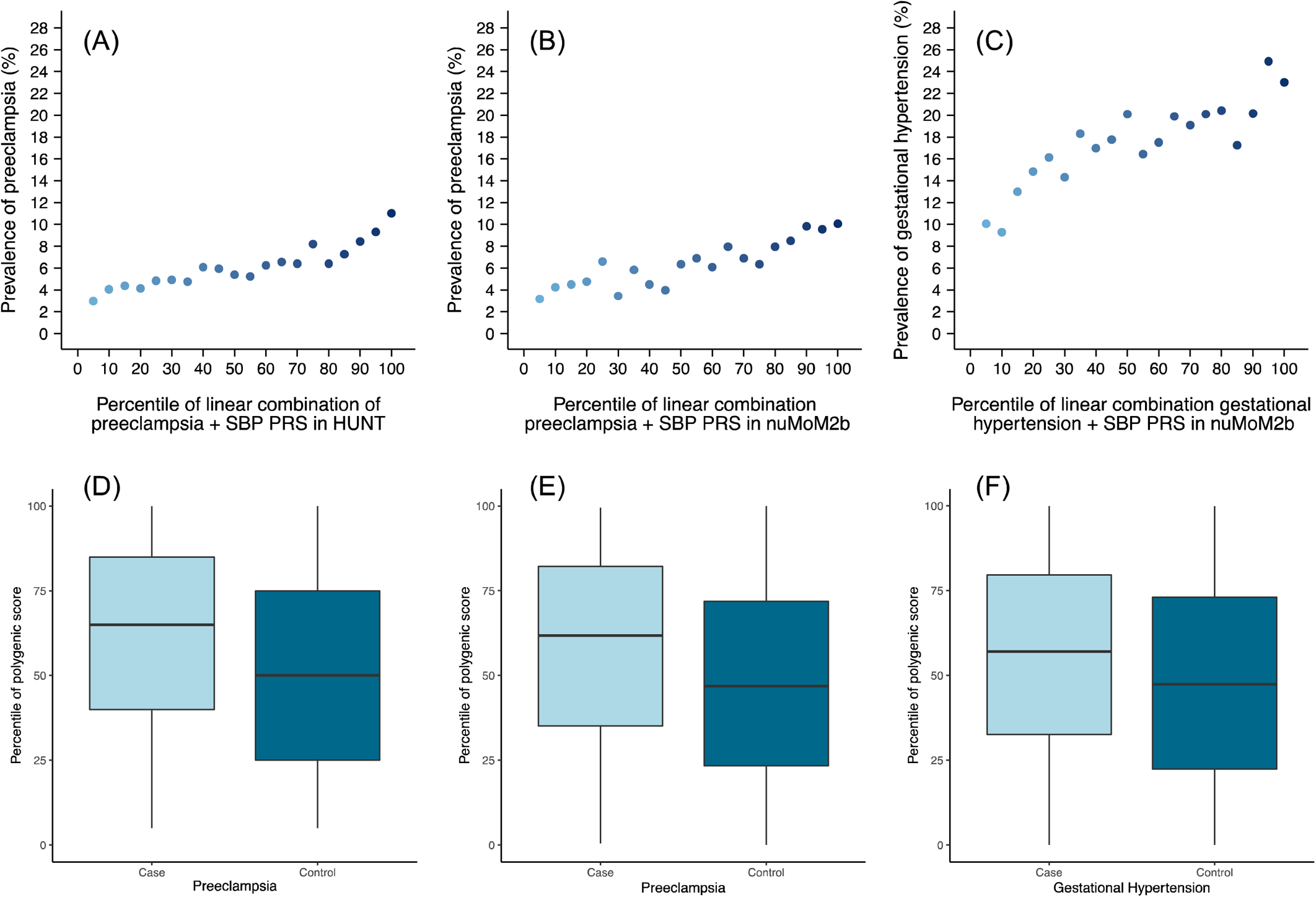
Polygenic prediction of preeclampsia/eclampsia and gestational hypertension in test cohorts. Polygenic risk scores (PRS) for preeclampsia/eclampsia and gestational hypertension were derived from our discovery genome-wide meta-analyses, tuned in the UK Biobank, and carried forward for testing in independent cohorts. Prevalence of preeclampsia/eclampsia vs. percentile of PRS_preeclampsia+SBP_ in **(A)** HUNT and **(B)** nuMoM2b. **(C)** Prevalence of gestational hypertension vs. percentile of PRS_GH+SBP_ in nuMoM2b. Distribution of PRS_preeclampsia+SBP_ by preeclampsia/eclampsia status in **(D)** HUNT and **(E)** nuMoM2b. **(F)** Distribution of PRS_GH+SBP_ by gestational hypertension status in nuMoM2b. Within each boxplot, horizontal lines reflect the median, top and bottom of the box reflect the interquartile range, and whiskers reflect the maximum and minimum value within each grouping.

We next tested PRS performance in the prospective, multi-ancestry nuMoM2b cohort of U.S. women recruited in the first trimester of their first pregnancy, including 481 (6.4%) who developed preeclampsia, 1,319 (17.5%) who developed gestational hypertension, and 5,744 with normotensive pregnancies (overall 73.6% European, 16.5% African, and 1.0% admixed American ancestry). Rates of preeclampsia/eclampsia ranged from ~4% among women in the bottom decile of PRS_preeclampsia+SBP_ to ~10% among the top decile of PRS_preeclampsia+SBP_ (**Fig. 2b**). Rates of gestational hypertension ranged from ~9% among women in the bottom decile of PRS_GH+SBP_ to ~24% among the top decile of PRS_GH+SBP_ (**Fig. 2c**). As in HUNT, incorporating SBP PRS in linear combination boosted PRS performance for HDPs, especially for gestational hypertension, although PRS performance was better for European ancestry vs. other women (**Supplementary Table 18**). After adjustment for age, PC 1-10, and self-reported race/ethnicity, PRS_preeclampsia+SBP_ and PRS_GH+SBP_ each predicted their respective outcomes (preeclampsia/eclampsia: OR 1.78, 95% CI 1.35-2.31, for top 10% vs. bottom 90% PRS_preeclampsia+SBP_, *P*=2.6×10^−5^; gestational hypertension: OR 1.52, 95% CI 1.26-1.84, for top 10% vs. bottom 90% PRS_GH+SBP_, *P*=1.0×10^−5^). As pre-pregnancy hypertension and obesity are established clinical predictors of HDPs, we performed further adjustment for first trimester SBP, antihypertensive medication use (as a marker of chronic hypertension), and body mass index (BMI). After this additional adjustment, the scores both remained predictive (preeclampsia/eclampsia: OR 1.64, 95% CI 1.23-2.15, for top 10% vs. bottom 90% PRS_preeclampsia+SBP_, *P*=5.1×10^−4^; gestational hypertension: OR 1.53, 95% CI 1.26-1.85, for top 10% vs. bottom 90% PRS_GH+SBP_, *P*=1.0×10^−5^). Compared with a model including age, PC 1-10, self-reported race/ethnicity, first trimester SBP, antihypertensive medication use, and first trimester BMI, addition of PRS_preeclampsia+SBP_ improved the C-statistic for preeclampsia/eclampsia from 0.690 to 0.701 (+0.011, 95% CI 0.001-0.021, Delong’s *P*=3.7×10^−2^). Similarly, addition of PRS_GH+SBP_ improved the C-statistic for gestational hypertension from 0.649 to 0.659 (+0.010, 95% CI 0.003-0.018, Delong’s *P*=5.7×10^−3^).

Low-dose aspirin starting after 12 weeks’ gestation represents an evidence-based but underutilized^42^ strategy to reduce risk of preeclampsia. To further probe the potential clinical impact of incorporating PRS to guide aspirin allocation, we examined aspirin eligibility according to current U.S. Preventive Service Task Force major criteria^43^ with and without addition of PRS_preeclampsia+SBP_ as an additional eligibility criterion in the nuMoM2b cohort. Among singleton, nulliparous women (i.e., the population enrolled in nuMoM2b), major criteria for aspirin eligibility are chronic pre-pregnancy hypertension, pre-gestational diabetes, kidney disease, and autoimmune disease.^43^ Major risk factors identified only a small proportion of the cohort that developed HDPs (**Table 3**), and aspirin would have been recommended in only 17.5% of women who developed preeclampsia/eclampsia (i.e., sensitivity of major risk factors alone was 17.5%), with a positive predictive value of 12.8%. Incorporating the top 10% of PRS_preeclampsia+SBP_ increased identification of the aspirin-eligible proportion to 30.4% of women with preeclampsia/eclampsia (i.e., sensitivity 30.4% [95% CI 26.2-34.5%]) with specificity of 83.3% (95% CI 82.5-84.2%), positive predictive value of 11.0% (95% CI 9.3-12.7%), and negative predictive value of 94.6% (95% CI 94.1-95.2%; **Table 3**). Expanding aspirin eligibility further to include the top 25% of PRS_preeclampsia+SBP_ captured nearly half (47.0%) of women who developed preeclampsia/eclampsia. Addition of high PRS_preeclampsia+SBP_ to major risk factors to up-classify risk of preeclampsia/eclampsia vs. other pregnancies yielded net reclassification of +1.8% (95% CI −0.3 to +4.0%) for top 5% PRS_preeclampsia+SBP_, +4.3% (95% CI 1.3-7.3%) for top 10% PRS_preeclampsia+SBP_, and +8.3% (95% CI 3.9-12.6%) for top 25% PRS_preeclampsia+SBP_ (**Table 3**).

**Table 3.**
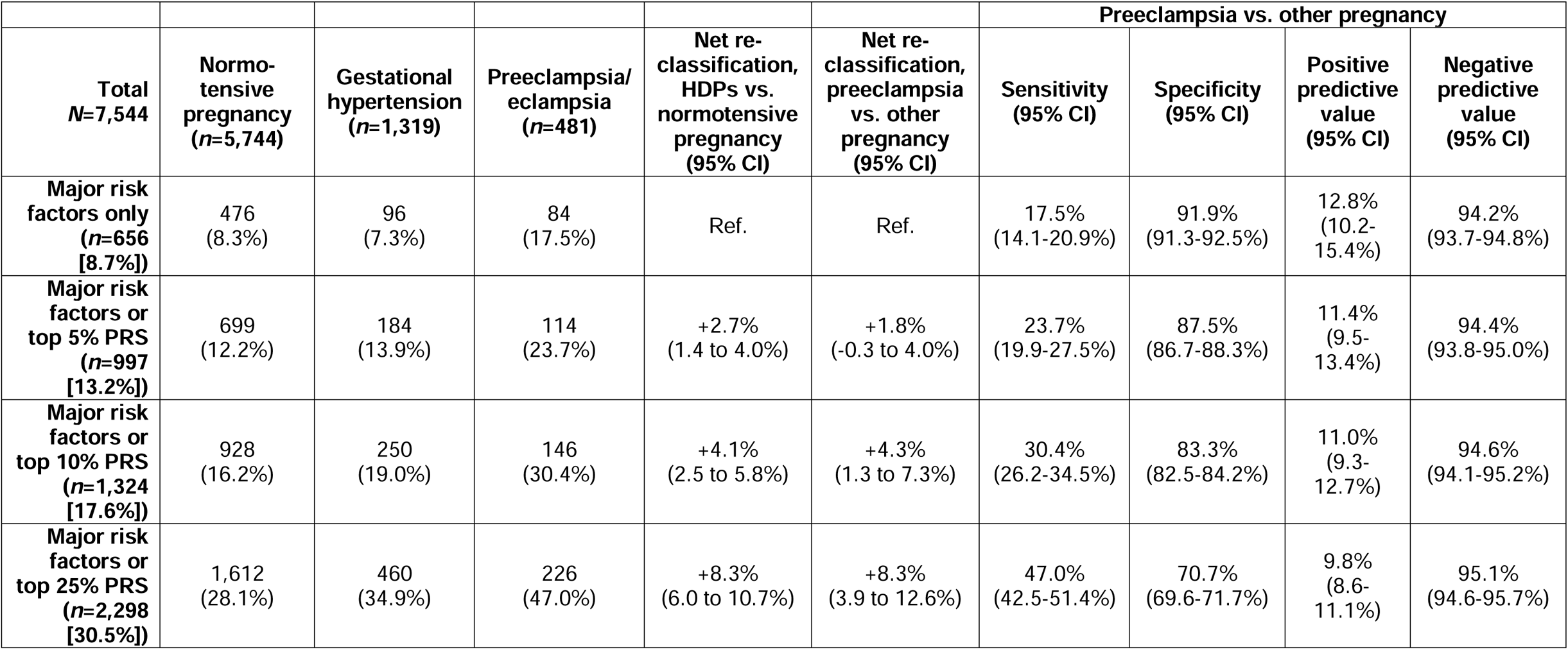
Aspirin eligibility to prevent preeclampsia using major clinical risk factors and polygenic risk. This Table reports the proportion of participants in nuMoM2b (nulliparous women with singleton gestations recruited in the first trimester of pregnancy) who experienced each pregnancy outcome in the index pregnancy (normotensive pregnancy, gestational hypertension, or preeclampsia/eclampsia) who would have been identified as candidates for low-dose aspirin to prevent preeclampsia according to U.S. Preventive Services Task Force major criteria, with or without addition of preeclampsia/eclampsia and systolic blood pressure combination polygenic risk score (PRS_preeclampsia+SBP_) as an additional criterion for aspirin eligibility. Net reclassification analyses reflect use of PRS_preeclampsia+SBP_ to up-classify risk for composite hypertensive disorders of pregnancy (HDPs) vs. normotensive pregnancy and for preeclampsia/eclampsia vs. other pregnancies when added to major risk factors.

### Phenome-wide associations with preeclampsia/eclampsia and gestational hypertension polygenic risk

We performed sex-stratified phenome-wide association analysis (pheWAS) for PRS_preeclampsia_ and PRS_GH_ across 1,445 phecode-based phenotypes in the UK Biobank. PRS_preeclampsia_ was significantly associated with 36 phenotypes in women and 37 phenotypes in men with Bonferroni-corrected statistical significance (*P*<0.05/1,445 = 3.5×10^−5^); PRS_GH_ was significantly associated with 25 and 32 phenotypes in women and men, respectively (**Fig. 3**). PRS_preeclampsia_ and PRS_GH_ were most strongly associated with hypertension in both sexes (PRS_preeclampsia_: OR_women_ 1.15 per SD, 95% CI 1.14-1.16, *P*=1.4×10^−175^; OR_men_ 1.12 per SD, 95% CI 1.11-1.13, P=7.5×10^−112^; PRS_GH_: OR_women_ 1.15 per SD, 95% CI 1.14-1.16, P=5.1×10^−184^; OR_men_ 1.13 per SD, 95% CI 1.11-1.14, P=1.3×10^−133^). Other strong phenotypic associations included hypercholesterolemia, type 2 diabetes, obesity, and atherosclerotic cardiovascular disease (**Supplementary Tables 19-20**). PRS_preeclampsia_ predicted ischemic heart disease in women (OR 1.09 per SD, 95% CI 1.07-1.11, *P*=4.4×10^−27^) and men (OR 1.09 per SD, 95% CI 1.07-1.10, *P*=2.6×10^−40^), as did PRS_GH_ (OR_women_ 1.07 per SD, 95% CI 1.05-1.08, *P*=4.4×10^−16^; OR_men_ 1.08 per SD, 95% CI 1.07-1.09, *P*=1.4×10^−35^). These similar associations between sexes suggest that most genes identified are not pregnancy-specific, but rather that pregnancy likely unmasks underlying risk. PRS_preeclampsia_ was also associated with several autoimmune phenotypes, including celiac disease, type 1 diabetes, hypothyroidism (in women), and a suggestive association with rheumatoid arthritis in women (*P*=5.7×10^−6^), whereas type 1 diabetes and celiac disease were not significantly associated with PRS_GH_ in either sex.

**Figure 3.**
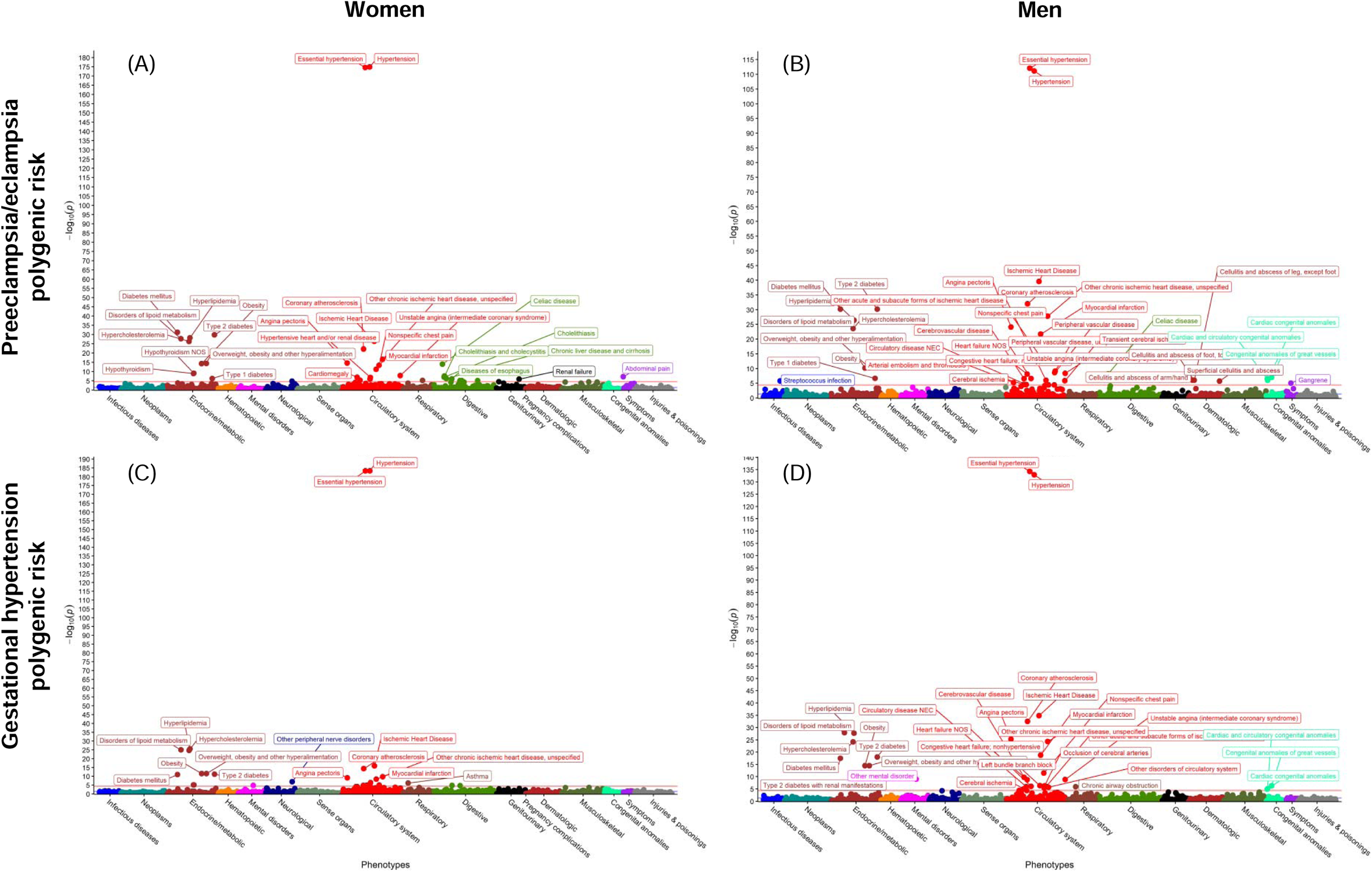
Sex-stratified phenome-wide association study of preeclampsia/eclampsia and gestational hypertension polygenic risk in the UK Biobank. Preeclampsia/eclampsia polygenic risk (A-B) and gestational hypertension polygenic risk (C-D) were associated with 1,445 phenotypes among women (A, C) and men (B, D) in the UK Biobank. Associations with phenotypes were tested using logistic regression with adjustment for age and PC 1-5.

## Discussion

We present an expanded multi-ancestry maternal genome-wide association analysis of preeclampsia/eclampsia and, to our knowledge, the first distinct GWAS of gestational hypertension, two pregnancy complications on the spectrum of HDPs with substantial associated public health impact. Altogether, we identified 18 independent genomic loci associated with preeclampsia/eclampsia and/or gestational hypertension: 13 loci associated with preeclampsia/eclampsia; 7 loci associated with gestational hypertension, of which 4 overlap with preeclampsia/eclampsia; and 2 additional loci associated with both phenotypes in multi-trait analysis. Identified loci highlight the role of angiogenesis and endothelial cell function (*FLT1*, *ZBTB46*), natriuretic peptide signaling (*NPPA*, *NPR3*, *FURIN*), renal glomerular function (*TRPC6*, *TNS2*, *PLCE1*), and immune dysregulation (*MICA*, *SH2B3*) in the pathogenesis of these conditions, with some loci (*FLT1*^16,17,36^, *WNT3A*^38,44^) previously described to influence risk via the fetal genome. Furthermore, we demonstrated that an HDP PRS augmented by a systolic blood pressure PRS stratified risk of HDP. Polygenic risk scores predicted HDP risk among nulliparous women independent of first trimester blood pressure and BMI, indicating potential clinical utility of these scores risk for pregnancy risk stratification. Collectively, these findings may have implications for advancing HDP prediction, prevention, and treatment.

First, our findings provide insights into mechanisms of HDP pathogenesis and underscore the causal role of blood pressure. As in previous GWAS of preeclampsia^17^, most genetic associations we identified were also associated with blood pressure^29^, and genetic correlation between blood pressure and the HDPs, especially gestational hypertension, was high. These findings align with prior work demonstrating heightened polygenic BP risk in those with HDPs^17,19,20^. Indeed, we observed that integrating SBP PRS with HDP PRS in linear combination further improved predictive performance for both preeclampsia/eclampsia and gestational hypertension. The recently published randomized CHAP trial of treatment for mild chronic hypertension in pregnancy demonstrated that lowering blood pressure pharmacologically reduced risk of progression to preeclampsia^45^, supporting the notion that elevated blood pressure is not merely a clinical manifestation of the HDPs but also plays a causal role in disease pathogenesis.

Second, our GWAS findings suggest a key role of natriuretic peptide signaling in the pathogenesis of the HDPs. The natriuretic peptides (e.g., ANP, B-type natriuretic peptide [BNP]) promote renal sodium excretion and counteract renin-angiotensin and sympathetic nervous system activation^46^. The lead variant at the *MTHFR-CLCN6* locus colocalized with *NPPA* expression in atrial tissue, its primary site of expression. ANP (the protein product of *NPPA*) also plays a role in uterine decidualization and spiral artery remodeling^47^, a process known to be impaired in the early pathogenesis of preeclampsia^1,10^. Furthermore, ANP is cleared from the circulation by the protein product of *NPR3*^48^, which was identified in the GWAS of gestational hypertension. Human data also support accelerated ANP clearance in the setting of preeclampsia^49^. Of note, our lead risk variant at the *MTHFR-CLCN6* locus has been previously associated with reduced levels of circulating N-terminal pro-BNP^50^. A recent analysis from the nuMoM2b cohort found that first trimester levels of N-terminal pro-BNP were unexpectedly lower among women who subsequently developed HDPs later in pregnancy after adjustment for race and BMI^51^. Collectively, these findings suggest that a relative deficiency in endogenous natriuretic peptide signaling may predispose to HDPs. Synthetic natriuretic peptides have been developed previously (e.g., nesiritide), and the natriuretic peptides may represent a future therapeutic target for direct or indirect modulation toward HDP prevention and/or treatment.

Third, our findings suggest several other potential novel mechanisms underlying HDP risk. Although *ZNF831* has been previously associated with hypertension, the mechanisms by which risk variants increase blood pressure have been unclear. In our conditional analysis at the *ZNF831* locus, we detected a second independent association with *ZBTB46*, which was also strongly colocalized with the lead *ZNF831* variant in multiple tissues, including arterial tissue. ZBTB46 is a transcription factor expressed in dendritic cells^52,53^ and in vascular endothelial cells, consistent with our observed expression in aortic endothelial cells, and *ZBTB46* overexpression suppresses endothelial cell proliferation and angiogenesis *in vitro*^54^. Furthermore, ZBTB46 is sensitive to shear stress^54^, which may have relevance to the hyperdynamic hemodynamic state of pregnancy.

In addition, we identified and replicated a novel association in the intergenic region between *PGR* and *TRPC6* (rs2508372), which has several plausible mechanistic links to the HDPs. Along with other newly identified HDP-associated loci (*TNS2*^55^ and *PLCE1*^29^), *TRPC6* has also been linked to glomerular function. It has been implicated in focal segmental glomerulosclerosis and diabetic nephropathy^56^ and mediates the proteinuria and renal dysfunction induced by exposure to hypertension and diabetes^57^. In addition, the progesterone receptor mediates the physiologic response to progesterone, including pregnancy-associated vasodilation^58^. As noted, *ARHGAP42* (adjacent to *PGR*), which was found to have reduced expression in preeclamptic placentas, regulates vascular tone and blood pressure^39^. Furthermore, this same intergenic lead variant has been associated with reduced circulating levels of matrix metalloproteinase-10^59^, which mediates vascular endothelial growth factor (VEGF)-induced angiogenesis^60^. Further research is necessary to clarify which of these potential mechanisms primarily mediate the preeclampsia/eclampsia risk associated with the *PGR/TRPC6* locus.

Fourth, associations at *MICA* and *SH2B3* highlight the role of immune function in preeclampsia pathogenesis. Differences in T-cell phenotypes and circulating pro- and anti-inflammatory cytokines in preeclampsia are well described^9,10^. We detected an association near *MICA* (encoding major histocompatibility complex class I polypeptide-related sequence A) within the *HLA* region, which has also been linked to multiple autoimmune diseases and may reflect the importance of maternal immune tolerance of fetal cells at the maternal-fetal interface in early placentation^10^. Indeed, we found that preeclampsia/eclampsia PRS predicted type 1 diabetes, celiac disease, and hypothyroidism. Subclinical thyroid dysfunction has been associated with increased risk for preeclampsia^61^, and previous Finnish data suggested term preeclampsia predicts incident hypothyroidism later in life^62^. SH2B3 (also known as LNK) is expressed primarily in endothelial and hematopoietic cells and negatively regulates cytokine signaling; reduced SH2B3 function has been linked to atherosclerosis as well as several autoimmune diseases^63^. The lead variant in our preeclampsia/eclampsia GWAS at *SH2B3* is in linkage disequilibrium (*D*’=0.96, *R*^2^=0.91)^64^ with the well-described coronary artery disease risk allele at this locus (rs3184504)^65^. Our lead *SH2B3* variant was also associated with increased leukocyte, red cell blood, and platelet counts in the UK Biobank (nealelab.is/uk-biobank) and with heightened levels of vascular cell adhesion protein 1, platelet glycoprotein IB alpha chain, interleukin-2 receptor, and lymphocyte activation gene 3 protein, among other immune-related proteins^50^. Recent data indicate that reduced SH2B3 function promotes neutrophil extracellular trap formation, a process implicated in preeclampsia pathogenesis^66,67^, and arterial thrombosis^68^. This process was dependent on the presence of oxidized phospholipids^68^, which are increased in the setting of preeclampsia vs. normotensive pregnancy^69,70^.

Fifth, we observed that polygenic risk scores for preeclampsia/eclampsia and gestational hypertension were predictive in multiple external test cohorts and independent of first trimester risk indicators (blood pressure and BMI), with women in the top PRS decile demonstrating 65-85% greater risk of preeclampsia/eclampsia vs. other women. Predictive accuracy of clinical risk factors for HDPs is modest^71,72^. Meta-analysis indicates that personal history of preeclampsia, pre-pregnancy chronic hypertension, and pre-pregnancy diabetes are associated with relative risks of 8.4, 5.1, and 3.7, respectively, for development of preeclampsia. However, nulliparity carries the largest population attributable fraction of associated risk for preeclampsia (approximately one-third)^12^, and most affected women lack any overt pre-pregnancy risk factors other than nulliparity^6^. Low-dose aspirin after 12 weeks’ gestation represents one evidence-based strategy to mitigate risk of preeclampsia and preterm birth in women at increased risk for preeclampsia^43^. Improving pregnancy risk prediction, especially in the first pregnancy, therefore remains a pressing clinical need to optimize HDP prevention^43,73^. First trimester screening algorithms have been developed, with the UK Fetal Medicine Foundation combined prediction model^74^ incorporating clinical factors, mean arterial pressure, uterine artery pulsatility index, and maternal serum pregnancy-associated plasma protein A and placental growth factor being most extensively validated to date^75^, although not currently endorsed by UK or US care guidelines^43^. Future studies are required to ascertain whether PRS may augment existing risk algorithms. In contrast with markers measured during pregnancy, PRS can be calculated anytime from birth, including preconception, and may therefore also inform preconception counseling and health optimization.

Although our GWAS included substantially more non-European individuals than prior GWAS, >80% of individuals were European, and as such, the preeclampsia/eclampsia and gestational hypertension PRS generally performed better in European ancestry women vs. others, consistent with many prior published PRS and a well-recognized challenge in contemporary genetics^76^. Ongoing efforts to include accurate, detailed pregnancy and reproductive history phenotypes in diverse genetic datasets and increase representation of non-European individuals will be critical to improve genetic discovery and cross-ancestry polygenic prediction and to achieve genomic equity^76,77^.

This study should be considered in the context of other limitations. Due to HDP phenotyping limitations in large datasets using ICD code-based ascertainment, some participants may have had preeclampsia superimposed on chronic hypertension rather than *de novo* preeclampsia, which may enrich genetic associations for hypertension predilection. In a subset of cohorts, however, the control group included women with chronic hypertension in pregnancy. Validation studies of ICD codes and registry diagnoses demonstrate that these approaches have modest sensitivity but high specificity and positive predictive value (>80%) in comparison with adjudicated HDP diagnoses^78–80^ and that accuracy may be higher for severe vs. mild preeclampsia^81^. We were unable to examine more granular HDP subtypes such as preeclampsia with severe features, HELLP (hemolysis, elevated liver enzymes, and low platelets) syndrome, preterm vs. term vs. postpartum onset, or HDP with vs. without accompanying fetal growth restriction. The underlying pathophysiology of the HDPs is heterogeneous and may vary across these subtypes; future adequately powered studies should examine these subtypes separately as implications for pregnancy care and long-term maternal health risk may differ^82^. In addition, we lacked paired maternal-fetal samples to condition maternal risk variants on fetal genotype, although other complementary analyses such as placental transcriptomics indicated particular variants more likely to be influencing risk via the fetal genome. Finally, snRNA-seq analyses were performed in male aortic tissue; while findings are consistent with current understanding of HDPs, future work is needed to verify that these results are consistent in women and determine whether additional insights may be apparent in women.

Overall, multi-ancestry genome-wide meta-analysis of preeclampsia/eclampsia and gestational hypertension revealed novel distinct and overlapping risk loci and enabled polygenic prediction of the HDPs, with implications for HDP prediction, prevention, and treatment.

## Methods

### Study cohorts, phenotypes, genotyping, and association analysis

Preeclampsia/eclampsia and gestational hypertension case and control counts and definitions for each cohort are summarized in Supplementary Tables 1-3. If an individual had qualifying codes for both preeclampsia/eclampsia and gestational hypertension, she was designated as having preeclampsia/eclampsia. In multi-ancestry cohorts, association analyses were performed within each ancestry group separately and subsequently meta-analyzed.

#### FinnGen

Cases were identified using ICD-8, -9, and -10 codes; controls were women in FinnGen without codes corresponding to hypertension and/or proteinuria during pregnancy (ICD-10 O10-O16). Sample genotyping in FinnGen was performed using Illumina (Illumina Inc., San Diego, CA) and Affymetrix (Thermo Fisher Scientific, Santa Clara, CA, USA) arrays. Genotype calls were made using GenCall or zCall for Illumina and AxiomGT1 algorithm for Affymetrix data as described previously^83,84^. Individuals were removed for ambiguous sex, genotype missingness >5%, heterozygosity >±4 SD, and non-Finnish ancestry. Variants were removed for missingness >2%, Hardy-Weinberg equilibrium (HWE) *P*<1×10^−6^, and minor allele count (MAC) <3. Pre-phasing was performed with Eagle v2.3.5 using 20,000 conditioning haplotypes. Genotypes were imputed with Beagle 4.1 using the population-specific Sequencing Initiative Suomi v3 imputation reference panel. Association analyses were performed using SAIGE^85^ with adjustment for age, genotyping batch, and PC 1-10.

#### Estonian Biobank

The Estonian Biobank is a population-based biobank with over 200,000 participants^86^. Preeclampsia/eclampsia cases were identified using ICD-10 O14-O15. Gestational hypertension cases were defined using ICD-10 O13. Controls were defined using ICD-10 O80-O84 and no codes for HDPs. All Estonian Biobank participants have been genotyped at the Core Genotyping Lab of the Institute of Genomics, University of Tartu, using the Illumina Global Screening Array v1.0, v2.0, and v2.0_EST arrays. Samples were genotyped and PLINK format files were created using Illumina GenomeStudio v2.0.4. Individuals were excluded from the analysis if their call-rate was <95% or if sex defined based on heterozygosity of X chromosome did not match sex in phenotype data. Before imputation, variants were filtered by call-rate <95%, HWE *P*<1×10^−4^, and MAF <1%. We also used the MAC filter --minMAC=5. Variant positions were updated to genome build 37 and all variants were changed to be from TOP strand using GSAMD-24v1-0_20011747_A1-b37.strand.RefAlt.zip files from https://www.well.ox.ac.uk/~wrayner/strand/. Pre-phasing was performed with Eagle v2.3 software using 20,000 conditioning haplotypes, and imputation was done using Beagle v.28Sep18.793 with effective population size n_e_=20,000. Population specific imputation reference of 2297 WGS samples was used. Analyses were carried out with SAIGE^85^, adjusting for year of birth and PC 1-10.

#### Genes & Health

Genes & Health is a cohort of British Pakistani and Bangladeshi individuals recruited primarily in East London, England^87^. Cases and parous controls were identified using qualifying ICD-10 and SNOMED codes (Supplementary Tables 1-2). Genotyping was performed using the Illumina Infinium Global Screening Array v3.0, and quality control was performed using Illumina GenomeStudio and PLINK v1.9. Individuals were removed who did not have Pakistani or Bangladeshi ancestry defined as >±3 SD from the mean of PC1 and those who self-reported another ethnicity. Variants with call rate <0.99, MAF <1%, and HWE *P*<1×10^−6^ were removed. Imputation was performed using the Michigan Imputation Server with the GenomeAsia reference panel. Association analyses were performed using SAIGE^85^ with adjustment for age, age^2^, and PC 1-10.

#### Michigan Genomics Initiative

The Michigan Genomics Initiative enrolls participants receiving care at Michigan Medicine and links biospecimen data to electronic health record (EHR) data. Preeclampsia cases were identified in the freeze 3 dataset using phecode 642.1 and its corresponding ICD codes^88^, and women lacking this phecode served as controls. Genotyping was performed using one of two versions of the Illumina Infinium CoreExome-24 bead array platform. Relatedness within the cohort was estimated using KING v2.1.3. Individuals were removed for discordant, missing, or ambiguous sex, kinship coefficient >0.45 with another participant, call rate <99%, estimated contamination >2.5%, or missingness on any chromosome >5%. Variants were excluded with poor intensity separation based on metrics from GenomeStudio (GenTrain score <0.15 or Cluster Separation score <0.3), overall call rate <99%, or HWE *P*-value <1×10^−4^. Genotypes were phased using EAGLE v2.4.1 and imputed using the TOPMed reference panel. Association analysis was conducted using SAIGE^85^ with adjustment for age, genotype array, and PC 1-10.

#### Mass General Brigham Biobank

The Mass General Brigham Biobank is a health system-based biobank linking genomic data to EHR data. Preeclampsia/eclampsia cases were identified using ICD-10 O14-O15, gestational hypertension cases were identified using ICD-10 O13, and controls were identified using ICD-10 O80-O82. Variants with MAF <1%, missingness per variant >1%, and HWE *P*-value <10^−6^ were removed. Imputation was performed using the TOPMed reference panel. Association analysis was performed with variants filtered by MAC ≥50 and INFO score ≥0.6 using REGENIE^89^ with adjustment for age, genotype batch, and PC 1-10.

#### Biobank Japan

Biobank Japan is a biobank of approximately 200,000 Japanese adults. Preeclampsia cases were identified using phecode 642 and its corresponding ICD codes, and other women constituted the control group^90^. Genotyping was performed using the Illumina HumanOmniExpressExome BeadChip or a combination of the Illumina HumanOmniExpress and HumanExome BeadChip. Individuals with call rates <98% or closely related individuals (PI_HAT >0.175 in PLINK) were excluded. Variants with call rate <99%, HWE *P*<1.0×10^−6^, and number of heterozygotes <5 were excluded. Genotype data were imputed with 1000 Genomes Project Phase 3 v5 genotype data and Japanese whole-genome sequencing data. Association analysis was performed using SAIGE^85^ with adjustment for age, age^2^, and PC 1-20.

#### BioMe

Bio*Me* is a health system-based biobank at the Icahn School of Medicine at Mount Sinai in New York, NY, USA. Preeclampsia/eclampsia and gestational hypertension cases were identified using ICD-10 codes O14-O15 and O13, respectively. Controls were other women enrolled in Bio*Me* who indicated a history of childbirth on their intake questionnaire. Genotyping was performed using the Illumina Global Screening Array. Individuals with ethnicity-specific heterozygosity rate that surpassed ±3 SD of the population-specific mean, those with a call rate of ≤95%, and those with discordance between EHR-recorded and genetic sex were removed. For variant-level quality control, sites with a call rate below 95% and sites with HWE *P*<1×10^−8^ were excluded. Imputation of variants was then performed with the Michigan Imputation Server pipeline using the TOPMed reference panel. Association analysis was performed separately in African, admixed American, and European ancestry women using SAIGE^85^ with adjustment for age, age^2^, and PC 1-10.

#### InterPregGen consortium

We incorporated summary statistics for preeclampsia from the discovery GWAS meta-analyses of European cohorts and Central Asian cohorts from the InterPregGen consortium^17^. European-ancestry discovery cohorts included GOPEC (United Kingdom), deCODE (Iceland), the Avon Longitudinal Study of Parents and Children (United Kingdom), MoBa (Norway), SSI (Denmark), and FINRISK (Finland) (7,219 cases and 155,620 controls). Central Asian cohorts included two Kazakh cohorts and one Uzbek cohort (2,296 cases and 2,059 controls). Cohort-specific preeclampsia and control definitions are summarized in Supplementary Table 3^17^. Fixed-effects inverse variance-weighted meta-analysis was performed in METAL^23^.

#### HUNT

The HUNT study is a population-based cohort study in Nord-Trøndelag County, Norway. Genotyped, parous, European-ancestry women were included in the present analysis. Preeclampsia/eclampsia was ascertained by linkage to the Medical Birth Registry of Norway, which defines preeclampsia as systolic blood pressure ≥140 mmHg and/or diastolic blood pressure ≥90 mmHg accompanied by proteinuria >0.3 g/24 h or >1+ on urine dipstick^91^. The current analysis includes genetic data from approximately 90% of participants from HUNT2 (1995-1997) and HUNT3 (2006-2008) who were genotyped by genome-wide SNP arrays in 2015^92,93^. Genotyping, quality control metrics, and imputation have been described previously^92^. Briefly, one of three different Illumina HumanCoreExome arrays (HumanCoreExome12 v1.0, HumanCoreExome12 v1.1, and UM HUNT Biobank v1.0) were used for genotyping the HUNT2 and HUNT3 samples^92^. Samples and variants with call rate <99% were excluded. Imputation was performed using 2,201 HUNT samples with whole genome sequencing, the Haplotype Reference Consortium, and TOPMed imputation panel (MAC >10). Association analysis was performed using SAIGE^85^ with adjustment for age, age^2^, and PC 1-10.

#### UK Biobank

The UK Biobank is a population-based cohort study of adult residents of the UK aged 40-69 years at the time of recruitment between 2006-2010. Preeclampsia/eclampsia and gestational hypertension cases were identified using qualifying ICD codes (Supplementary Table 1); controls were women self-reporting a history of at least one live birth at the time of study enrollment without qualifying ICD codes for preeclampsia/eclampsia or gestational hypertension or any self-reported history of HDPs. Genotyping was performed using the UK BiLEVE Axiom Array or the UK Biobank Axiom Array (both Affymetrix, Santa Clara, CA). Individuals with single nucleotide variant missingness ≥10% were excluded. Imputation was performed centrally using the Haplotype Reference Consortium, UK10K, and 1000 Genome reference panels^94^. Variants were required to pass the following quality control filters: MAF ≥1%, single nucleotide variant missingness <10%, and HWE *P*≥10^−15^, MAC ≥50, and INFO score ≥0.6. Association analysis was performed in European-ancestry participants using REGENIE^89^ with adjustment for age, genotyping array, and PC 1-10.

#### Penn Medicine Biobank (PMBB)

PMBB is a health system-based biobank at the University of Pennsylvania, Philadelphia, PA, USA. Preeclampsia/eclampsia cases were identified using ICD-10 codes O14-O15 or phecode 642.1. Gestational hypertension cases were identified using ICD-10 code O13. Controls were other women enrolled in PMBB. Genotyping was performed using the Illumina Global Sequencing Array v2.0; genotype data were imputed to the TOPMed reference panel using the Michigan Imputation Server. Variants with MAF<1%, missing rate >10%, and HWE *P*<10^−8^ were filtered from the GWAS. Association analysis was performed in REGENIE separately for African-ancestry and European-ancestry women with adjustment for age, age^2^, and PC 1-5.

#### nuMoM2b

The nuMoM2b study is a prospective U.S. pregnancy cohort of nulliparous women enrolled in the first trimester of pregnancy between 2012-2015. HDPs were determined by chart abstraction and adjudication according to previously published definitions^95^. Gestational hypertension was defined as SBP ≥140 mmHg or DBP ≥90 mmHg on two occasions ≥6 hours apart or one occasion with subsequent antihypertensive therapy after 20 weeks’ gestation, excluding blood pressures recorded during the second stage of labor, without other qualifying features for preeclampsia or eclampsia. Preeclampsia was defined according to the same blood pressure criteria plus proteinuria or other findings meeting criteria for severe features, including HELLP syndrome^95^. Women documented as meeting these same blood pressure criteria before 20 weeks’ gestation were designated as having chronic hypertension. All women were at risk for development of preeclampsia/eclampsia; only those without chronic hypertension were at risk for the outcome of gestational hypertension. Blood pressure and BMI were recorded at the first trimester study visit, which occurred at a mean (SD) 11.6 (1.5) weeks’ gestation. Genotyping was performed using the Illumina Infinium Multi-Ethnic Global D2 BeadChip. Individuals related within two degrees by the KING algorithm were removed. Variants with MAF <1%, genotyping rate <95%, and HWE *P*<5×10^−6^ were removed^96^. After phasing with EAGLE, imputation was performed for European, African, and admixed American ancestry subjects using the TOPMed reference panel via the TOPMed Imputation Server. Association analysis was performed in REGENIE separately by ancestry group (European, African, and admixed American) with adjustment for age and PC 1-10. The same women in nuMoM2b with imputed genotypes were used for external testing of optimized PRS (described below).

### Genome-wide meta-analysis and replication

Variants from GWAS summary statistics were matched by genome build 38 position and alleles. GWAS summary statistics that were in genome build 37 were lifted to genome build 38 using UCSC liftOver (https://genome.ucsc.edu/cgi-bin/hgLiftOver). We used METAL^23^ to perform fixed-effect inverse variance-weighted meta-analysis. Correction for genomic inflation factor was carried out prior to meta-analysis. Meta-analyses were conducted among discovery cohorts, among follow-up cohorts, and across all cohorts. Given the potential overlap of 400 preeclampsia/eclampsia cases from FINRISK between the InterPregGen meta-analysis and FinnGen (8.4% of FinnGen cases and 2.3% of overall discovery cases) and 7,805 controls (5.7% of FinnGen controls and 1.7% of overall discovery controls), we conducted a sensitivity analysis excluding FinnGen (Supplementary Table 4).

Lead variants for preeclampsia/eclampsia and gestational hypertension were interrogated in multi-ancestry meta-analysis of follow-up cohorts using METAL: HUNT (preeclampsia/eclampsia only), UK Biobank, PMBB (European- and African-ancestry women), and nuMoM2b (European-, African-, and admixed American-ancestry women). *P*<0.05 in follow-up cohorts, consistent direction of effect in follow-up cohorts, and genome-wide significance (*P*<5×10^−8^) in combined meta-analysis of discovery and follow-up cohorts indicated replication.

### Conditional and joint analysis

We conducted conditional analysis using GCTA-COJO^24^ on the multi-ancestry meta-analyses of preeclampsia/eclampsia and gestational hypertension to identify additional association signals at the genome-wide significant loci. We used the European LD reference from a randomly selected set of 10,000 unrelated individuals. The LD panel included variants with MAF > 1% and INFO ≥0.3. The analysis was restricted to variants within ±1 Mb from lead variants (*P*<5×10^−8^). In COJO, the lead variants were conditioned from each chromosome and independent variants were iteratively included. All variants were then simultaneously fitted in the joint analysis. Variants with *P*<2×10^−7^ were considered genome-wide significant. One additional variant with conditional *P*<2×10^−7^ for association with preeclampsia/eclampsia was identified on chromosome 20 (*P*=1.4×10^−8^).

### Genetic correlation

We used LD score regression (LDSC, https://github.com/bulik/ldsc)^25^ with pre-computed LD scores for 1.2 million HapMap3 variants after excluding MHC region in the European population (https://data.broadinstitute.org/alkesgroup/LDSCORE/eur_w_ld_chr.tar.bz2) to calculate genetic correlation between preeclampsia/eclampsia and gestational hypertension and correlation of each HDP with systolic and diastolic blood pressure. In addition, the LDSC intercept indicates potential confounding due to potential population stratification and cryptic relatedness. In combined meta-analysis of discovery and follow-up cohorts, we observed intercepts of 1.03 for preeclampsia/eclampsia and 0.95 for gestational hypertension.

### Colocalization analysis

We obtained the tissue-specific gene expression from GTEx data portal for 52 tissues^32^. We used marginal effect sizes, standard errors, and MAF for all SNPs within ±500 of lead variants from discovery analysis (*P*<5×10^−8^) as the input. We performed colocalization using the coloc.abf() function in R package coloc v4. The H_4_ test statistic estimates the posterior probability of a shared causal variant between preeclampsia/eclampsia or gestational hypertension and expression of a particular gene. H_4_ >0.7 indicated strong evidence of colocalization, H_4_ 0.5-<0.7 indicated weak evidence of colocalization, and H_4_ <0.5 indicated no colocalization.

### Polygenic prioritization of causal genes

We performed additional causal gene prioritization using the PoPS method^34^. Briefly, PoPS integrates GWAS summary statistics with gene expression, biology pathways, and predicted protein-protein interaction data to identify likely causal genes at genome-wide significant loci. A linear model was trained to predict gene-level association scores and estimate Z-scores indicating the confidence of the causal role at a given locus. In total, PoPS scores were calculated for 18,000 genes. The top 5 available prioritized genes within 500 kilobases of lead preeclampsia/eclampsia and gestational hypertension variants were extracted and compared with the results of other *in silico* analyses.

### Placental transcriptome data

Genes nearest to lead variants, colocalization hits, and genes prioritized within the top 5 by either Open Targets variant-to-gene score or PoPS score were queried in a publicly available database of placental gene expression (https://www.obgyn.cam.ac.uk/placentome/)^35^. Samples were obtained from a prospective cohort of nulliparous women in Cambridge, United Kingdom. Differential expression analysis for preeclampsia included 82 preeclampsia cases and 82 controls samples matched on presence of labor, Cesarean section, gestational age, fetal sex, smoking status, maternal BMI, and maternal age. RNA sequencing was performed on placental biopsy specimens with a median sequencing depth of 101 million reads per sample. Differential expression (in log_2_-fold change) and corresponding *P*-values were generated using DESeq2; *P*-values were then adjusted for multiple comparisons using the Benjamini-Hochberg method. We report differentially expressed prioritized genes with adjusted *P* <0.05 (Supplementary Table 14).

### Gene expression in human aortic tissue

We queried expression of prioritized genes in subpopulations of vascular smooth muscle cells, fibromyocytes, fibroblasts, endothelial cells, macrophages, natural killer T cells, and neuronal cells in a dataset of single-nuclei RNA sequencing from human aortic tissue. In total, 1,114 unique molecular identifiers were obtained per cell. Prioritized genes were nearest genes, genes with strong colocalization, genes with weak colocalization plus prioritization by another method (top 5 Open Targets variant-to-gene score or PoPS), and genes prioritized by another method plus statistically significant differential placental transcription in the human placental transcriptome browser^35^’; of these genes, 24 were available in the single-nuclei RNA sequencing dataset (Extended Data Fig. 3). Single-nuclei RNA sequencing was performed on non-atherosclerotic aortic root tissue from two individuals obtained during coronary artery bypass graft surgery. Both individuals contributing aortic tissue specimens were European-ancestry men, aged 49 and 51 years, with hypertension, hypercholesterolemia, and coronary artery disease. Both were using aspirin and a statin preoperatively. A total of 4,537 nuclei were obtained for downstream analysis. Cell types and subtypes were defined using top marker genes and pathway enrichment scores. Raw *Cellranger* output data were filtered for removal of ambient RNA using *cellbender* in ‘full’ running mode. The resultant filtered cell-gene matrix was used for quality control and downstream analysis. All preliminary quality-control and clustering were performed using *scanpy*. Any cells with fewer than 300 genes captured or greater than 0.1% mitochondrial reads were excluded from the analysis. Each sample was processed with *Scrublet* to exclude doublets. The top 10,000 variable genes were used for analysis. Relative expression of queried genes in each cell type against other cell types in normal aortic tissue was quantified as Z-scores.

### Derivation and testing of genome-wide polygenic risk scores

We used PRS-CS to derive genome-wide PRS for preeclampsia/eclampsia and gestational hypertension from the corresponding discovery GWAS summary statistics and for systolic blood pressure from the Million Veteran Program GWAS summary statistics^26^. PRS-CS is a Bayesian approach that estimates posterior effect sizes by placing a continuous shrinkage prior on variant effect sizes^41^. The preeclampsia/eclampsia PRS included 1,087,033 HapMap3 variants, the gestational hypertension PRS included 1,087,916 HapMap3 variants, and the SBP PRS included 1,064,898 HapMap3 variants. PRS were trained on the UK Biobank European LD panel. Individual-level polygenic scores were generated in the tuning and test datasets as the sum of genotypes*weights using PLINK. We used logistic regression to test the association of each PRS with preeclampsia/eclampsia and gestational hypertension with adjustment for age, age^2^, and PC 1-10. PRS were tuned in the UK Biobank. Specifically, we used a small-scale grid search of global shrinkage parameter values (1, 1×10^−2^, 1×10^−4^, and 1×10^−6^) for each PRS as recommended to identify the that produced the best predictive performance as measured by *R*^2^ in the tuning dataset. We then fitted a linear combination of optimized preeclampsia/eclampsia PRS and SBP PRS for the outcome of preeclampsia/eclampsia and a linear combination of gestational hypertension PRS and SBP PRS for the outcome of gestational hypertension. The optimal linear combination derived for preeclampsia/eclampsia was 0.1889\**Z*_preeclampsia/eclampsia_ + 0.1864\**Z*_SBP_, and the linear combination derived for gestational hypertension was 0.1662\**Z*_gestational_hypertension_ + 0.3050\**Z*_SBP_. We then carried these weighed linear combination scores forward for final testing in nuMoM2b (European, African, and admixed American ancestry groups) and HUNT (European ancestry). PRS performance was evaluated using the OR for top decile vs. bottom 90% of PRS, the OR per SD of PRS, and Nagelkerke’s *R*^2^.

We tested whether PRS correctly re-classified nuMoM2b participants with HDPs as aspirin-eligible in comparison with the major criteria endorsed by the U.S. Preventive Services Task Force^43^. Major criteria include: history of preeclampsia, which does not apply in nuMoM2b as all women were nulliparous; multifetal gestation, which does not apply in nuMoM2b as all women had singleton pregnancies; chronic hypertension, defined as a diagnosis of hypertension before pregnancy or blood pressure ≥140/90 mmHg on two occasions at least 6 hours apart prior to 20 weeks’ gestation; pre-gestational diabetes type 1 or type 2; any pre-pregnancy kidney disease; and autoimmune disease, defined here as antiphospholipid antibody syndrome, systemic lupus erythematosus, rheumatoid arthritis, inflammatory bowel disease (Crohn’s disease or ulcerative colitis), or “other collagen vascular or autoimmune disease.” We calculated sensitivity, specificity, positive predictive value, and negative predictive value for major risk factors with or without different thresholds for PRS_preeclampsia+SBP_ for the prediction of preeclampsia/eclampsia, as well as net reclassification for composite HDPs vs. normotensive pregnancy and for preeclampsia/eclampsia vs. all other pregnancies. Confidence intervals for sensitivity, specificity, positive predictive value, and negative predictive value were calculated using the normal approximation. Given use of PRS to up-classify risk, net reclassification was calculated as *P*(up|case) - *P*(up|non-case). Bootstrap resampling performed 1,000 times was used to estimate 95% confidence intervals for net reclassification.

### Phenome-wide association analysis

We tested the association of preeclampsia/eclampsia PRS and gestational hypertension PRS with 1,445 phecode-based combined prevalent and incident phenotypes^97^ in sex-stratified fashion among genotyped UK Biobank participants with adjustment for age and PC 1-5 using the PheWAS package^98^ in R 3.6.0 (https://github.com/PheWAS/PheWAS). Bonferroni-corrected *P*<0.05/1,445 = 3.5×10^−5^ indicated statistical significance.

### Ethics approval

FinnGen was approved by the Coordinating Ethics Committee of the Helsinki and Uusimaa Hospital District. The Estonian Committee on Bioethics gave ethical approval for the work conducted in the Estonian Biobank. The South East Research Ethics Committee gave ethical approval for the work conducted in Genes & Health. The University of Michigan Medical School Institutional Review Board gave ethical approval for the analyses conducted in the Michigan Genomics Initiative. The Mass General Brigham Institutional Review Board gave ethical approval for the work conducted in the Mass General Brigham Biobank. Biobank Japan received ethics approval from the Institute of Medical Science, the University of Tokyo, the RIKEN Yokohama Institute, and all participating hospitals. Bio*Me* received ethics approval from the Icahn School of Medicine at Mt. Sinai Institutional Review Board. We used publicly available summary statistics for discovery GWAS from the InterPregGen consortium; all contributing studies received ethics approval as reported previously^17^. The work conducted in HUNT was approved by the Regional Committee for Ethics in Medical Research, Norway (2018/2488). The Penn Medicine Biobank received ethics approval from the University of Pennsylvania Institutional Review Board. The North West Multi-centre Research Ethics Committee approved the UK Biobank; the Mass General Brigham Institutional Review Board approved secondary data analyses of the UK Biobank (application #7089). The nuMoM2b study was approved by the institutional review boards of each participating site. All participants in all studies contributing data for this study signed informed consent for participation and the use of data in research.

## Data Availability

GWAS summary statistics for preeclampsia/eclampsia and gestational hypertension and corresponding genome-wide polygenic scores will be made publicly available upon publication. Summary statistics used in this meta-analysis are publicly available for FinnGen r6 (https://www.finngen.fi/en/access_results) and for BioBank Japan (https://pheweb.jp/pheno/PreEclampsia). Summary statistics from the InterPregGen consortium are available at https://ega-archive.org. Placental transcriptome data are publicly available at https://www.obgyn.cam.ac.uk/placentome/.

## Data availability

Upon publication, GWAS summary statistics for preeclampsia/eclampsia and gestational hypertension will be made publicly available through the Broad Institute CVDi portal, and corresponding genome-wide polygenic scores will be posted to the PGS Catalog. Summary statistics used in this meta-analysis are publicly available for FinnGen r6 (https://www.finngen.fi/en/access_results) and for BioBank Japan (https://pheweb.jp/pheno/PreEclampsia). Summary statistics from the InterPregGen consortium are available at https://ega-archive.org. Placental transcriptome data are publicly available at https://www.obgyn.cam.ac.uk/placentome/.

## Code availability

The code used to conduct these analyses is available at https://github.com/buutrg/HDP

## Acknowledgements

This work was supported by grants from the U.S. National Heart Lung and Blood Institute K08HL166687 (M.C.H.), K08HL146963 (K.J.G.), R01 HL163234 (R.S., K.J.G.), R01HL139865 (R.D.), R01HL155915 (R.D.), DP2HL152423 (R.M.G.), U01HL166060 (R.M.G.), R03HL148483 (R.M.G.), R01HL142711 (P.N.), R01HL127564 (P.N.), R01HL148050 (P.N.), R01HL151283 (P.N.), R01HL148565 (P.N.), R01HL135242 (P.N.), R01HL151152 (P.N.); the American Heart Association 940166 (M.C.H.), 979465 (M.C.H.); the Korea Health Industry Development Institute HI19C1330 (S.M.J.C.); Harvard Catalyst Medical Research Investigator Training Program (A.P.P.); National Human Genome Research Institute U01HG011719 (A.P.P. and P.N.); the Belgian American Educational Foundation (A.S.); the U.S. National Institute of General Medical Sciences R35GM147197 (R.F.G.), R35GM124836 (R.D.); National Institute of Diabetes and Digestive and Kidney Diseases R01DK125782 (P.N.); National Institute of Child Health and Human Development R01HD101246 (D.M.H.); Preeclampsia Foundation (K.J.G., R.S.); Fondation Leducq TNE-18CVD04 (P.N.); and the Massachusetts General Hospital Paul and Phyllis Fireman Endowed Chair in Vascular Medicine (P.N.).

The work was conducted under UK Biobank application 7089. The Trøndelag Health Study (HUNT) is a collaboration between HUNT Research Centre (Faculty of Medicine and Health Sciences, NTNU, Norwegian University of Science and Technology), Trøndelag County Council, Central Norway Regional Health Authority, and the Norwegian Institute of Public Health. The genotyping in HUNT was financed by the National Institutes of Health; University of Michigan; the Research Council of Norway; the Liaison Committee for Education, Research and Innovation in Central Norway; and the Joint Research Committee between St Olavs hospital and the Faculty of Medicine and Health Sciences, NTNU. The genetic investigations of the HUNT Study is a collaboration between researchers from the K.G. Jebsen Center for Genetic Epidemiology, NTNU and the University of Michigan Medical School and the University of Michigan School of Public Health. The K.G. Jebsen Center for Genetic Epidemiology is financed by Stiftelsen Kristian Gerhard Jebsen; Faculty of Medicine and Health Sciences, NTNU, Norway. DNA extraction and genotyping of nuMoM2b was supported by the Indiana University Precision Health Initiative. The authors thanks other members of the Estonian Biobank research team: Lili Milani, Andres Metspalu, Tõnu Esko, Mari Nelis, Reedik Mägi, and Georgi Hudjashov. Genes & Health is/has recently been core-funded by Wellcome (WT102627, WT210561), the Medical Research Council (UK) (M009017, MR/X009777/1, MR/X009920/1), Higher Education Funding Council for England Catalyst, Barts Charity (845/1796), Health Data Research UK (for London substantive site), and research delivery support from the NHS National Institute for Health Research Clinical Research Network (North Thames). Genes & Health is/has recently been funded by Alnylam Pharmaceuticals, Genomics PLC; and a Life Sciences Industry Consortium of Astra Zeneca PLC, Bristol-Myers Squibb Company, GlaxoSmithKline Research and Development Limited, Maze Therapeutics Inc, Merck Sharp & Dohme LLC, Novo Nordisk A/S, Pfizer Inc, Takeda Development Centre Americas Inc. The authors thank Social Action for Health, Centre of The Cell, members of our Community Advisory Group, and staff who have recruited and collected data from volunteers. We thank the NIHR National Biosample Centre (UK Biocentre), the Social Genetic & Developmental Psychiatry Centre (King’s College London), Wellcome Sanger Institute, and Broad Institute for sample processing, genotyping, sequencing and variant annotation. We thank: Barts Health NHS Trust, NHS Clinical Commissioning Groups (City and Hackney, Waltham Forest, Tower Hamlets, Newham, Redbridge, Havering, Barking and Dagenham), East London NHS Foundation Trust, Bradford Teaching Hospitals NHS Foundation Trust, Public Health England (especially David Wyllie), Discovery Data Service/Endeavour Health Charitable Trust (especially David Stables), NHS Digital for GDPR-compliant data sharing backed by individual written informed consent. Most of all, we thank all of the volunteers participating in Genes & Health. Current Genes & Health Research Team (in alphabetical order by surname): Shaheen Akhtar, Mohammad Anwar, Elena Arciero, Omar Asgar, Samina Ashraf, Saeed Bibi, Gerome Breen, Raymond Chung, Charles J Curtis, Shabana Chaudhary, Maharun Chowdhury, Grainne Colligan, Panos Deloukas, Ceri Durham, Faiza Durrani, Fabiola Eto, Sarah Finer, Joseph Gafton, Ana Angel Garcia, Chris Griffiths, Joanne Harvey, Teng Heng, Sam Hodgson, Qin Qin Huang, Matt Hurles, Karen A Hunt, Shapna Hussain, Kamrul Islam, Vivek Iyer, Ben Jacobs, Ahsan Khan, Cath Lavery, Sang Hyuck Lee, Robin Lerner, Daniel MacArthur, Daniel Malawsky, Hilary Martin, Dan Mason, Rohini Mathur, Mohammed Bodrul Mazid, John McDermott, Shefa Miah, Caroline Morton, Bill Newman, Elizabeth Owor, Asma Qureshi, Samiha Rahman, Nishat Safa, Miriam Samuel, John Solly, Daniel Stow, Farah Tahmasebi, Richard C Trembath, Karen Tricker, Nasir Uddin, David A van Heel, Klaudia Walter, Caroline Winckley, John Wright, Julia Zollner.

## Ethics declarations

M.C.H. reports consulting fees from CRISPR Therapeutics, advisory board service for Miga Health, and grant support from Genentech, all unrelated to this work. K.J.G. has served as a consultant for BillionToOne, Aetion, and Roche for projects unrelated to this work. R.S. is a co-founder of Magnet Biomedicine, unrelated to this work. R.D. reports receiving grants from AstraZeneca, grants and nonfinancial support from Goldfinch Bio, being a scientific co-founder, consultant, and equity holder for Pensieve Health (pending), and being a consultant for Variant Bio, all unrelated to this work. P.N. reports grant support from Amgen, Apple, AstraZeneca, Boston Scientific, and Novartis, spousal employment and equity at Vertex, consulting income from Apple, AstraZeneca, Novartis, Genentech / Roche, Blackstone Life Sciences, Foresite Labs, and TenSixteen Bio, and is a scientific advisor board member and shareholder of TenSixteen Bio and geneXwell, all unrelated to this work.

## Extended Data

**Extended Data Figure 1.**
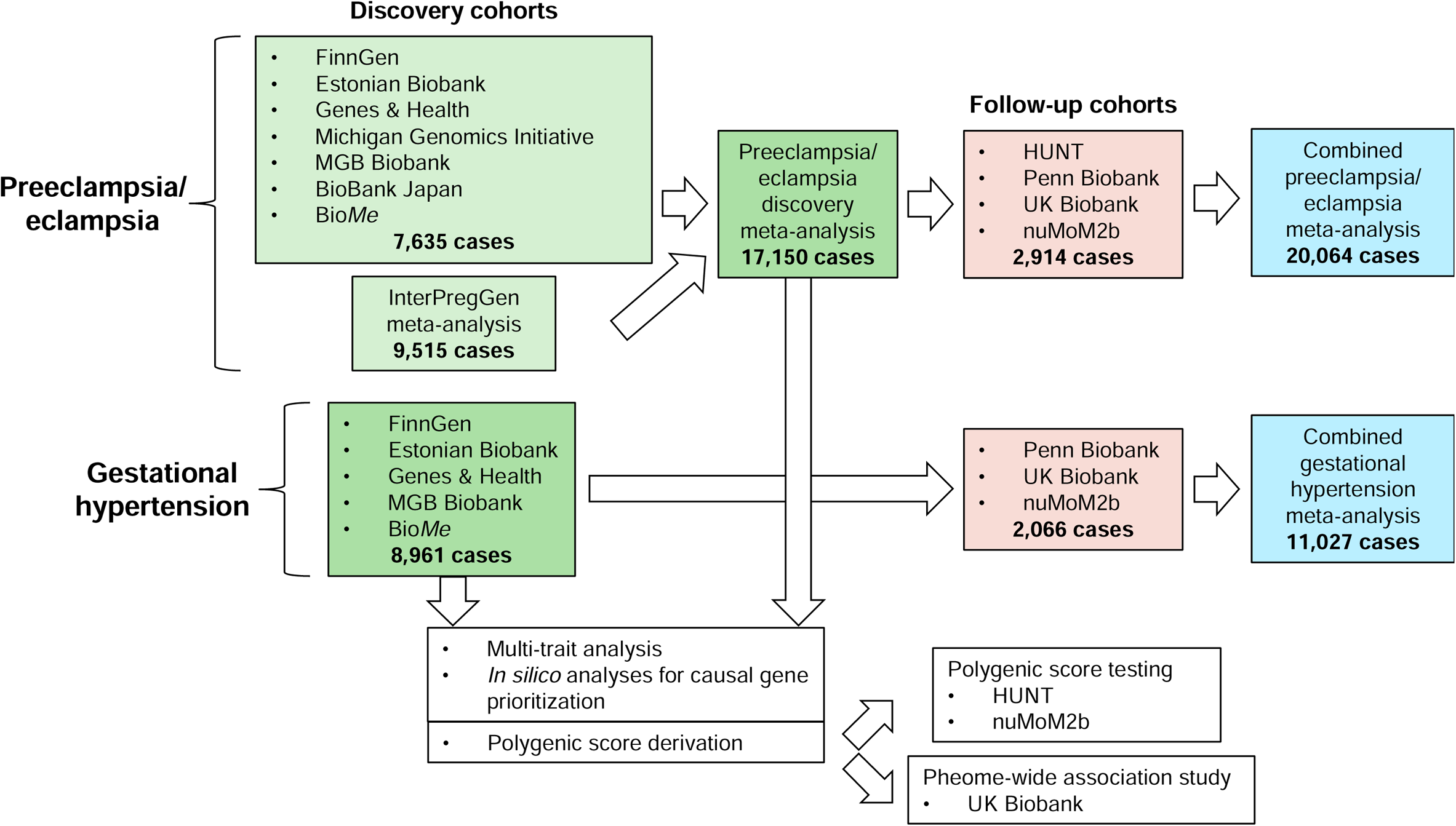
Flow chart summarizing the study design and contributing cohorts.

**Extended Data Figure 2.**
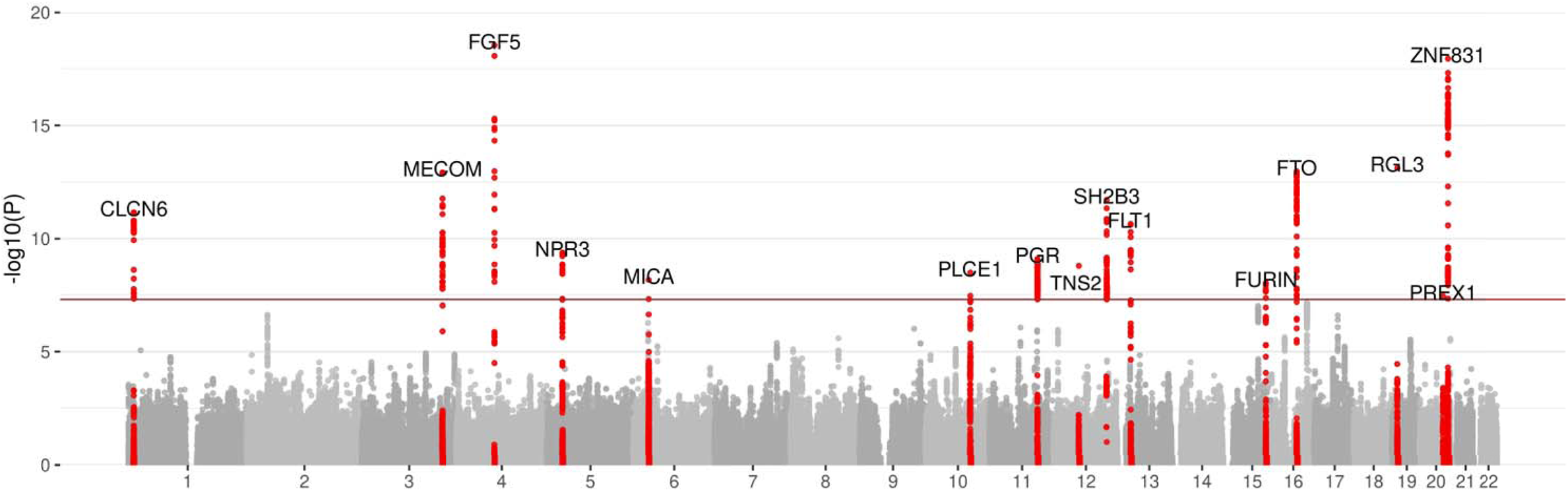
Results of multi-trait analysis of genome-wide summary statistics (MTAG) for preeclampsia/eclampsia from joint analysis of summary statistics for preeclampsia/eclampsia and gestational hypertension in discovery cohorts. The plot displays chromosomal position on the X-axis and −log(10) of the P-value on the Y-axis.

**Extended Data Figure 3.**
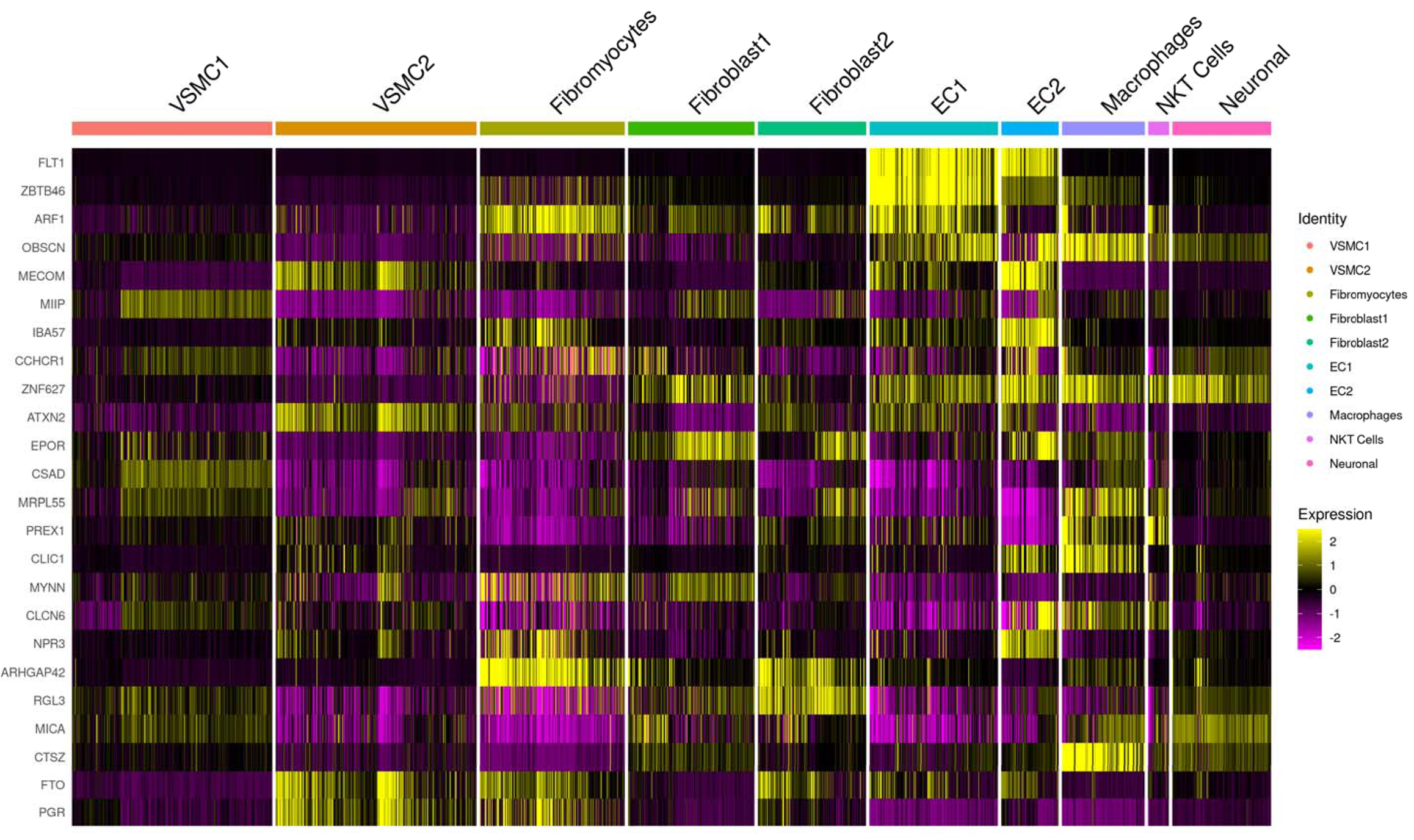
Relative expression of prioritized genes in human aortic cells with single-nuclei RNA sequencing. We analyzed expression of genes prioritized by genome-wide meta-analysis of preeclampsia/eclampsia and gestational hypertension and secondary *in silico* analyses in a dataset of single-nuclei RNA sequencing from two normal human flash-frozen aortic specimens. Most prioritized genes were enriched in endothelial cell populations and/or macrophages.

