## Supplementary Figures for "Genome-wide meta-analysis identifies novel maternal risk variants and enables polygenic prediction of preeclampsia and gestational hypertension"

**Supplementary Figure 1.** Quantile-quantile plots for combined meta-analysis of discovery and follow-up cohorts for: (A) Preeclampsia/eclampsia; (B) Gestational hypertension.

**Supplementary Figure 2.** Manhattan plots of preeclampsia/eclampsia and gestational hypertension in discovery cohorts.

**Supplementary Figure 1. Quantile-quantile plots for combined meta-analysis of discovery and follow-up cohorts for: (A) Preeclampsia/eclampsia; (B) Gestational hypertension.**

(A)

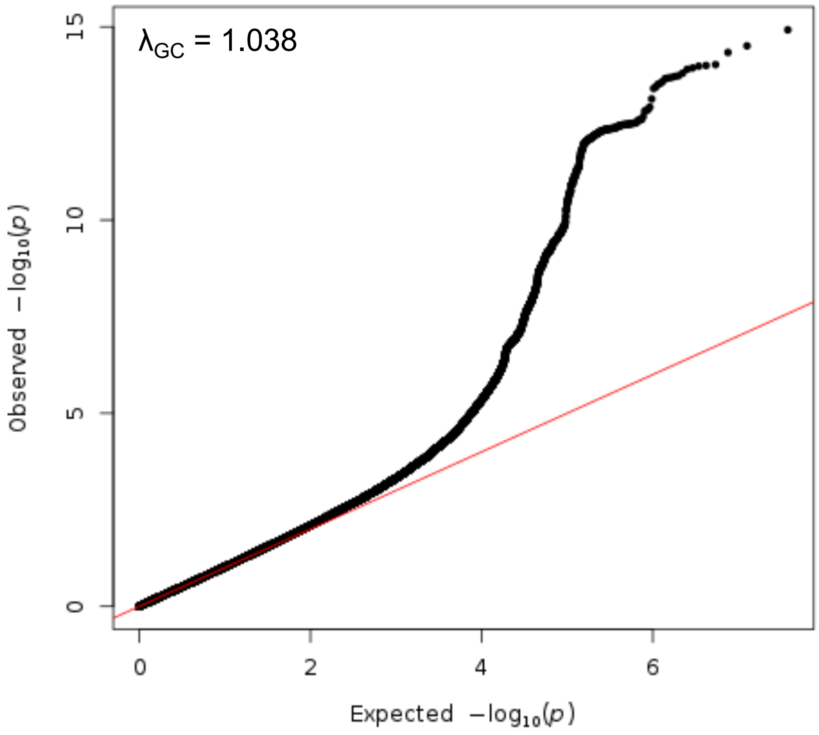


(B)

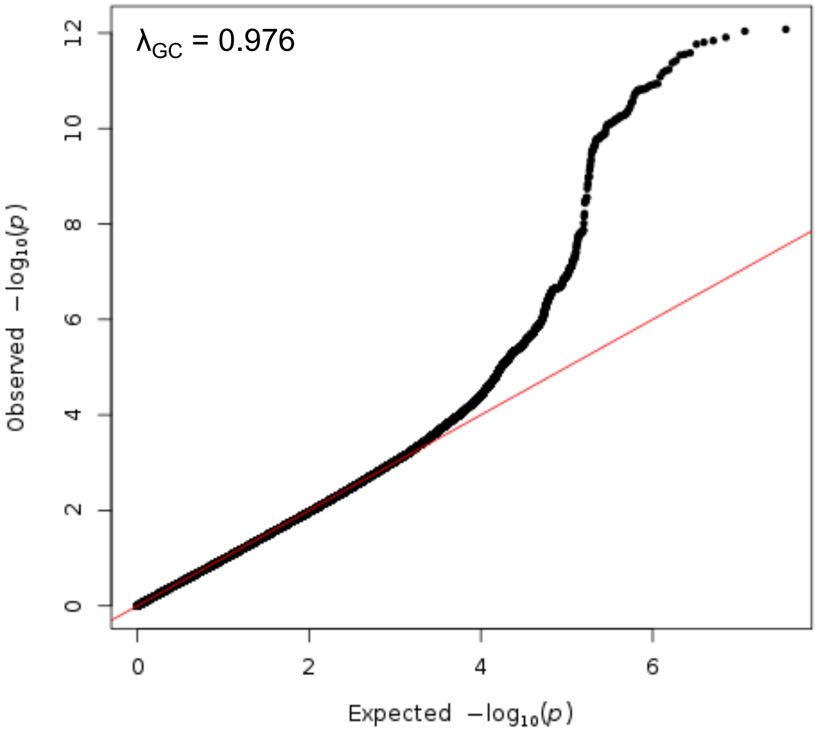

**Supplementary Figure 2. Manhattan plots of preeclampsia/eclampsia and gestational hypertension in discovery cohorts.** Manhattan plots (chromosomal position on the X-axis and -log(10) of the P-value on the Y-axis) are displayed for **(A)** preeclampsia/eclampsia in 17,150 cases and 451,241 controls and **(B)** gestational hypertension in 8,961 cases and 184,925 controls. Analyses included multi-ancestry meta-analysis of common variants (minor allele frequency ≥1%). Loci are labeled by the gene nearest to the lead variant.

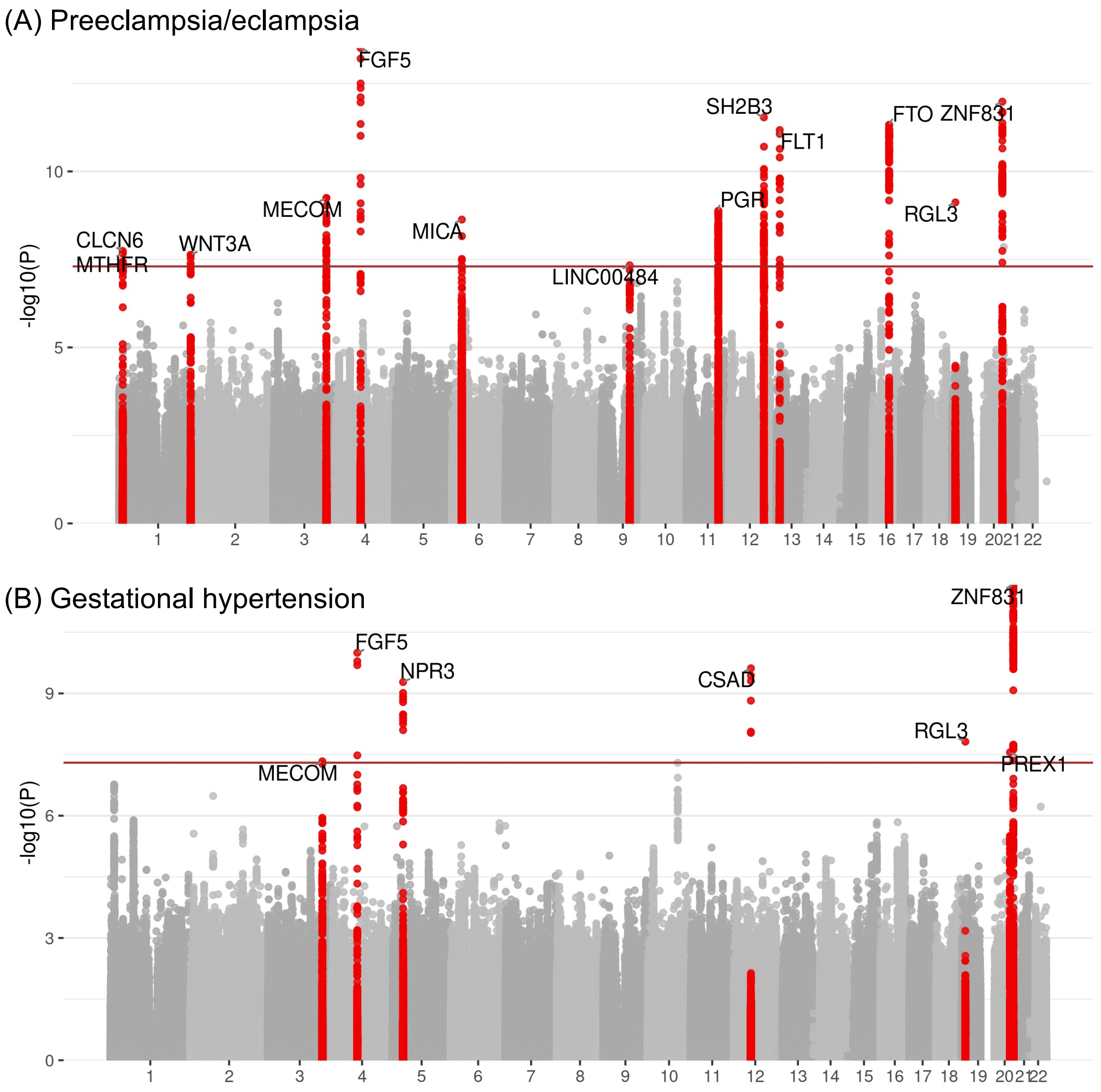
